# Pregnant women’s attitudes and behaviours towards antenatal vaccination against Influenza and COVID-19 in the Liverpool City Region, United Kingdom: cross-sectional survey

**DOI:** 10.1101/2022.09.13.22279846

**Authors:** Samantha Kilada, Neil French, Elizabeth Perkins, Dan Hungerford

**Affiliations:** Department of Clinical Infection Microbiology and Immunology, Institute of Infection, Veterinary & Ecological Sciences, University of Liverpool, Liverpool, UK; Centre for Global Vaccine Research, University of Liverpool, Liverpool, UK; Institute of Population Health, University of Liverpool, Liverpool, UK

**Keywords:** SARS-CoV-2, COVID-19, pregnancy, influenza, vaccine, attitudes, behaviours, survey, cross-sectional

## Abstract

**Objective:** Influenza poses a serious health risk to pregnant women and their babies. Despite this risk, influenza vaccine uptake in pregnant women in the UK is less than 50%. Little is known about how COVID-19 affects pregnant women, but its management may affect attitudes and behaviours towards vaccination in pregnancy. The study objectives were to establish attitudes and knowledge of pregnant women towards influenza disease and influenza vaccination and to compare these to attitudes and knowledge about COVID-19 and COVID-19 vaccination.

**Design:** A cross-sectional survey was conducted using an online questionnaire distributed through local advertisement and social media outlets. Information was sought on attitudes and knowledge of influenza and COVID-19 and their respective vaccines.

**Participants and setting:** Pregnant women residing in Liverpool City Region, UK

**Results:** Of the 237 respondents, 73.8% reported receiving an influenza vaccine. Over half (56.5%) perceived themselves to be at risk from influenza, 70.5% believed that if they got influenza, their baby would get ill, and 64.6% believed getting influenza could hurt their baby, 60.3% believed that the influenza vaccine would prevent their baby from getting ill, and 70.8% believed it would protect their baby. Only 32.9% of respondents stated they would receive the COVID-19 vaccine if it were available to them. However, 80.2% stated they would receive a COVID-19 vaccine if they were not pregnant. Most of the women stated that they would accept a vaccine if recommended to them by healthcare professionals.

**Conclusions:** Acceptance of the influenza and COVID-19 vaccines during pregnancy seems to be more related to the safety of the baby rather than the mother. Women perceived their child to be more at risk than themselves. Information about influenza and COVID-19 vaccine safety as well as healthcare provider recommendations play an important role in vaccine uptake in pregnant women.

**Strengths and limitations of this study:**

- The study provides information on how a pandemic affects vaccine attitudes and behaviours during pregnancy.
- The study compares and contrasts attitudes and behaviours towards influenza and COVID-19 vaccines.
- The study provides new information relating to barriers to COVID-19 vaccine acceptance and provides insights into mechanisms for improving uptake.
- The sample size is small and self-selected which might lead to an over-representation of women likely to accept or have strong opinions on vaccinations.
- Responses to the questions on vaccine status are self-reported, not provided from healthcare records.

## Introduction

Influenza illness is a serious risk to health in pregnant women, especially for those who are in a “high-risk” category, meaning those with underlying health conditions. After the H1N1 swine influenza epidemic in 2009, it was recommended that pregnant women in the UK receive the influenza vaccine[1]. However, uptake of the influenza vaccine in pregnant women is consistently below 50% and remained low in the UK at 43.5% for the period between September 2020 and February 2021[2].

Due to immunological and physiological changes associated with pregnancy, pregnant women are at risk of more severe side effects following influenza infection[3]. These adverse side effects include increased risk of miscarriage, premature birth, lowered female growth rates, and increased rate of maternal morbidity and mortality[4-6].

Inactivated influenza vaccines have an excellent and well characterised safety profile and can be given at any point during the gestational period with the benefits of vaccine extending to both the high-risk pregnant mother and the infant[7]. Influenza vaccines have been shown to be safe for pregnant women and have no associations with premature birth, low birth weight, or respiratory issues requiring ventilation at birth in infants[8]. Further, evidence suggests transplacental transport of antibodies following maternal vaccination which are protective for the baby. Giving the influenza vaccine to pregnant women is very effective in preventing lab-confirmed cases of influenza in their infants up to six months of age[9]. A US-based study found that when women were vaccinated against influenza during pregnancy, there was an 81% decrease in influenza hospitalizations in their babies within the first six months after birth[10].

Since the COVID-19 pandemic began, there has been limited research at this time regarding COVID-19 and pregnant women. Due to immunological and physiological changes in pregnancy, pregnant women may, in theory, be more susceptible to COVID-19 infection[11]. COVID-19 is a respiratory illness caused by the SARS-CoV-2 virus, a highly pathogenic human coronavirus[12]. Respiratory illness can be known to adversely affect the foetus as low oxygen levels in the mother can lead to foetal compromise[13].

A multinational cohort study involving 18 countries (including the UK) found that pregnant women who tested positive for COVID-19 had higher rates of adverse side effects including maternal mortality and preterm births compared to non-infected women[14]. A study from Texas, USA found that neonatal infection was 3% and these infants were born to asymptomatic or mildly asymptomatic mothers[15]. There is also a possible link found between infection with SARS-CoV2 in the third trimester of pregnancy and progressive coagulopathy[16].

On the 16^th^ of April, 2021, the Joint Committee on Vaccination and Immunisation (JCVI), UK recommended the use of COVID-19 vaccines for pregnant women in line with the age group specific roll out; prior to this recommendation, the vaccine was not recommended to pregnant women[17]. As of October 2021, almost 20% of critically ill patients with COVID-19 are unvaccinated pregnant women[18]. In response, the NHS has urgently encouraged pregnant women to get the COVID-19 vaccine[18]. Due to its recent recommendation, there is a dearth of evidence from clinical trials or from reported adverse events from COVID-19 vaccines in pregnant women and clinical trials are now increasingly unlikely to deliver rigorous data due to limited recruitment opportunities. As the vaccine has become available to pregnant women in the United Kingdom, it is important to understand attitudes towards receiving the vaccine in order to inform communication measures to support uptake of the vaccine in the population of pregnant women.

We conducted a cross-sectional survey of the attitudes and behaviours of pregnant women in the Liverpool City Region, UK towards influenza illness, COVID-19, and towards antenatal vaccination against influenza and COVID-19. This survey is one part of a larger project regarding vaccine attitudes and behaviours in pregnancy. We explored some of the factors that influence pregnant women’s attitudes towards vaccines and how these attitudes affect vaccine hesitancy or acceptance.

## Methods

### Population and setting

This study only included women who were currently pregnant and living in the Liverpool City Region, Merseyside (Liverpool, Knowsley, Sefton, St. Helens, Wirral, Halton) in the North West of the United Kingdom. The most recent statistics list 15,045 live births in Merseyside for 2019 and 15,632 for the year prior[19]. In the region, influenza vaccine uptake was 40.5% for September 2020 to January 2021[2].

### Data collection

A questionnaire was developed using JISC (Joint Information Systems Committee) online surveys and was live from 30 October 2020 through 30 April 2021 (Supplementary file 1). This questionnaire was developed using previously created questionnaires as a basis and adding topic-specific questions as they would aid in answering the main question of the study. Additional questions were added if they were relevant to pregnancy of health behaviours during pregnancy. A summary of the study’s purpose, inclusion criteria, confidentiality, and right to withdraw was presented on the first page of the survey prior to obtaining informed valid consent. The survey included questions examining pregnant women’s health behaviours, such as participation in exercise, the use of antenatal vitamins, and whether or not they smoke. It also included Likert scale questions about the respondent perceptions of illness severity for both influenza and COVID-19 and, were they to contract either, their perceptions of the risks of their own infection on their child as well as their perceptions of the potential risk to others. Questions were also asked about attitudes and beliefs about the influenza vaccine, COVID-19 vaccine, and vaccines in general to understand the factors that lead to vaccine acceptance or hesitancy. As the pertussis vaccination is recommended during pregnancy between 16 and 32 weeks, status of receiving this vaccination was asked[20]. Demographic details were collected on ethnic group, age, occupation, and socioeconomic status. All questions asked in the questionnaire, with the exception of giving consent, were optional and, thus, women could choose to not answer some of the questions. Participants were also asked if they wished to enter a prize draw at the end of the questionnaire for the chance to win a £100 Amazon voucher; this was optional.

A photo advertisement was created for the study and the link to the questionnaire was provided via multiple social media outlets (Supplementary file 2, Image S1). Social media was used while businesses were closed during the national lockdown in the UK and flyers were used once businesses opened again. It was shared via Twitter as well as multiple Facebook groups, including pregnant mother groups in the Liverpool City Region, antenatal class pages, city council pages, charity groups, and community centres. Social media pages were chosen through recommendation by PPI panel and colleagues as well as through search for groups in the area. Flyers were created with a QR code and distributed among local shops, community centres, and places of worship in the area.

### Patient and Public Involvement

The Institute of Infection, Veterinary, & Ecological Sciences at the University of Liverpool has a patient and public involvement and engagement (PPIE) group which provides an opportunity for discussion about influenza vaccine research. PPIE members had identified reducing inequalities in influenza vaccine uptake as a policy priority. The PPIE panel was used throughout the research process to review study processes and tools. The Liverpool Babies PPIE Group also assisted in the recruitment of the sample by distributing study details and information through their social media and contacts. The findings from this study will be shared through PPIE panels and with maternal and public health services.

### Data analysis

Descriptive analysis was carried out using R version 4.0.3 (R Core Team, Vienna, Austria) and RStudio (Supplementary file 3). We excluded from the analysis any respondents who did not meet the strict eligibility and inclusion criteria, this criteria being women who are currently pregnant and live in the Liverpool City Region. In the R code, an upper limit was 75 was used to analyse the age question as participants over this age were likely not currently pregnant and therefore don’t fit the inclusion criteria.

For the ease of understanding, the Likert scale questions were recoded so that responses of “Strongly Disagree” and “Disagree” were relabelled as “Disagree,” and responses of “Strongly Agree” and “Agree” were relabelled as “Agree.” These questions were analysed and correlated with the demographic questions. Due to the sample size questions were collapsed for analysis. Also, due to the sample size, statistical analysis was not appropriate.

Income was recoded so that responses of “<£10,000,” “£10,001-20,000,” and “£20,001-30,000” were recoded as “≤£30,000.” Occupations were recoded and put into groups of “More advantaged,” “Less advantaged,” or exceptions (full-time students who in the NS-SEC are not classified in the aforementioned groups) based on National Statistics Socio-economic classification (NS-SEC)[21]. The category of “Ethnically Diverse” was recoded to include the following ethnicities: Mixed/multiple ethnic groups (White and Asian, White and Black African, White and Black Caribbean, Other), Asian British/Asian (Chinese, Pakistani, Indian, Bangladeshi, Other), Black British/Black/African/Caribbean, and Other ethnic groups (Arab, Other).

## Results

### Demographics

The total number of survey responses was 252, and, of these, 237 (94.0%) fulfilled the inclusion criteria. Of those excluded, 3 (1.2%) were age outliers (elderly or the respondent did not complete their age details) and 12 (4.8%) lived outside of the study region. The age distribution of respondents was from 20 to 43 years old, (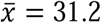 years old; Table 1). Most respondents were over 20 weeks pregnant (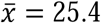 weeks). Two-hundred-fourteen of the 237 responses (90.3%) were completed before the recommendation of the COVID-19 vaccine for pregnant women in the UK in April 2021.

**Table 1.** Demographics for questionnaire respondents in relation to those who were vaccinated/unvaccinated against influenza and those who were accepting, undecided, or against the possibility of the COVID-19 vaccine.

| **Demographic Variables** | **Overall**<br>**N=237 (100%)** | **Vaccinated/**<br>**intend to**<br>**against**<br>**Influenza**<br>**N=196 (100%)** | **Unvaccinated**<br>**against**<br>**Influenza**<br>**N=39 (100%)** | **Would have COVID-**<br>**19 Vaccine**<br>**N=78 (100%)** | **Undecided**<br>**about having**<br>**COVID-19**<br>**Vaccine**<br>**N=38 (100%)** | **Would not**<br>**have COVID-19**<br>**Vaccine**<br>**N=121 (100%)** |
| --- | --- | --- | --- | --- | --- | --- |
| **Age** | **N=237** | **N=196** | **N=39** | **N=78** | **N=38** | **N=121** |
| Median [Min, Max] | 31.0 [20.0, 43.0] | 32.0 [20.0, 43.0] | 30.0 [21.0, 42.0] | 31.5 [21.0, 43.0] | 32.0 [20.0, 42.0] | 31.0 [20.0, 42.0] |
| Q1, Q3 | 28.0, 35.0 | 28.0, 35.0 | 27.0, 33.5 | 27.3, 35.0 | 29.0, 34.0 | 38.0, 34.0 |
| **Weeks Pregnant** | **N=235** | **N=194** | **N=39** | **N=77** | **N=38** | **N=120** |
| Median [Min, Max] | 27.0 [0, 41.0] | 28.0 [0, 41.0] | 20.0 [0, 37.0] | 30.0 [6.00, 40.0] | 27.5 [0, 39.0] | 24.5 [0, 41.0] |
| Q1, Q3 | 19.0, 33.0 | 21.0, 34.0 | 15.0, 29.5 | 21.0, 35.0 | 19.8, 33.0 | 18.0, 32.0 |
| **Occupation** | **N=237** | **N=196** | **N=39** | **N=78** | **N=38** | **N=121** |
| More advantaged groups NS-SEC groups 1-4 | 189 (79.7%) | 161 (82.1%) | 27.0 (69.2%) | 64.0 (82.1%) | 28.0 (73.7%) | 97.0 (80.2%) |
| Less advantaged groups NS-SEC groups 5-8 | 43.0 (18.1%) | 32.0 (16.3%) | 11.0 (28.2%) | 10.0 (12.8%) | 10.0 (26.3%) | 23.0 (19.0%) |
| Exceptions | 5.00 (2.1%) | 3.00 (1.5%) | 1.00 (2.6%) | 4.00 (5.1%) | 0 (0%) | 1.00 (0.8%) |
| **Income** | **N=234** | **N=193** | **N=39** | **N=77** | **N=38** | **N=119** |
| ≤£30,000 | 42.0 (17.9%) | 31.0 (16.1%) | 10.0 (25.6%) | 10.0 (13.0%) | 7.00 (18.4%) | 25.0 (21.0%) |
| £30,001-60,000 | 110 (47.0%) | 93.0 (48.2%) | 16.0 (41.0%) | 31.0 (40.3%) | 21.0 (55.3%) | 58.0 (48.7%) |
| >£60,000 | 82.0 (35.0%) | 69.0 (35.7%) | 13.0 (33.3%) | 36.0 (46.8%) | 10.0 (26.3%) | 36.0 (30.3%) |
| **Education** | **N=237** | **N=196** | **N=39** | **N=78** | **N=38** | **N=121** |
| GCSE or similar | 15.0 (6.3%) | 8.00 (4.1%) | 7.00 (17.9%) | 2.00 (2.6%) | 4.00 (10.5%) | 9.00 (7.4%) |
| NVQ or similar | 15.0 (6.3%) | 12.0 (6.1%) | 3.00 (7.7%) | 5.00 (6.4%) | 2.00 (5.3%) | 8.00 (6.6%) |
| A-level or similar | 40.0 (16.9%) | 34.0 (17.3%) | 6.00 (15.4%) | 11.0 (14.1%) | 9.00 (23.7%) | 20.0 (16.5%) |
| Undergraduate | 66.0 (27.8%) | 55.0 (28.1%) | 10.0 (25.6%) | 24.0 (30.8%) | 12.0 (31.6%) | 30.0 (24.8%) |
| Postgraduate | 100 (42.2%) | 86.0 (43.9%) | 13.0 (33.3%) | 35.0 (44.9%) | 11.0 (28.9%) | 54.0 (44.6%) |
| Other | 1.00 (0.4%) | 1.00 (0.5%) | 0 (0%) | 1.00 (1.3%) | 0 (0%) | 0 (0%) |
| **Ethnicity** | **N=237** | **N=196** | **N=39** | **N=78** | **N=38** | **N=121** |
| White British | 215 (90.7%) | 177 (90.3%) | 36.0 (92.3%) | 73.0 (93.6%) | 33.0 (86.8%) | 109 (90.1%) |
| White Other | 12.0 (5.1%) | 10.0 (5.1%) | 2.00 (5.1%) | 2.00 (2.6%) | 4.00 (10.5%) | 6.00 (5.0%) |
| Ethnically Diverse | 10.0 (4.2%) | 9.00 (4.6%) | 1.00 (2.6%) | 3.00 (3.8%) | 1.00 (2.6%) | 6.00 (5.0%) |
| **Number of Children** | **N=237** | **N=196** | **N=39** | **N=78** | **N=38** | **N=121** |
| This is my first | 114 (48.1%) | 93.0 (47.4%) | 20.0 (51.3%) | 35.0 (44.9%) | 19.0 (50.0%) | 60.0 (49.6%) |
| 1 | 96.0 (40.5%) | 81.0 (41.3%) | 14.0 (35.9%) | 36.0 (46.2%) | 15.0 (39.5%) | 45.0 (37.2%) |
| 2 | 22.0 (9.3%) | 18.0 (9.2%) | 4.00 (10.3%) | 7.00 (9.0%) | 2.00 (5.3%) | 13.0 (10.7%) |
| 3+ | 5.00 (2.1%) | 4.00 (2.0%) | 1.00 (2.6%) | 0 (0%) | 2.00 (5.3%) | 3.00 (2.5%) |
| **Number in Household** | **N=235** | **N=194** | **N=39** | **N=78** | **N=38** | **N=119** |
| 1 | 7.00 (3.0%) | 6.00 (3.1%) | 1.00 (2.6%) | 1.00 (1.3%) | 2.00 (5.3%) | 4.00 (3.4%) |
| 2 | 107 (45.5%) | 88.0 (45.4%) | 18.0 (46.2%) | 36.0 (46.2%) | 14.0 (36.8%) | 57.0 (47.9%) |
| 3 | 92.0 (39.1%) | 77.0 (39.7%) | 14.0 (35.9%) | 33.0 (42.3%) | 16.0 (42.1%) | 43.0 (36.1%) |
| 4+ | 29.0 (12.3%) | 23.0 (11.9%) | 6.00 (15.4%) | 8.00 (10.3%) | 6.00 (15.8%) | 15.0 (12.6%) |
| **Smartphone** | **N=235** | **N=194** | **N=39** | **N=77** | **N=37** | **N=121** |
| Yes | 235 (100.0%) | 194 (100.0%) | 39.0 (100.0%) | 77.0 (100.0%) | 37.0 (100.0%) | 121 (100.0%) |
| **Responses by Month** | **N=237** | **N=196** | **N=39** | **N=78** | **N=38** | **N=121** |
| November 2020 | 21.0 (8.9%) | 20.0 (10.2%) | 1.00 (2.6%) | 9.00 (11.5%) | 3.00 (7.9%) | 9.00 (7.4%) |
| December 2020 | 13.0 (5.5%) | 9.00 (4.6%) | 4.00 (10.3%) | 4.00 (5.1%) | 1.00 (2.6%) | 8.00 (6.6%) |
| January 2021 | 63.0 (26.6%) | 57.0 (29.1%) | 5.00 (12.8%) | 23.0 (29.5%) | 9.00 (23.7%) | 31.0 (25.6%) |
| February 2021 | 85.0 (35.9%) | 70.0 (35.7%) | 14.0 (35.9%) | 23.0 (29.5%) | 15.0 (39.5%) | 47.0 (38.8%) |
| March 2021 | 30.0 (12.7%) | 22.0 (11.2%) | 8.00 (20.5%) | 9.00 (11.5%) | 6.00 (15.8%) | 15.0 (12.4%) |
| April 2021 | 25.0 (10.5%) | 18.0 (9.2%) | 7.00 (17.9%) | 10.0 (12.8%) | 4.00 (10.5%) | 11.0 (9.1%) |
| **Before/After COVID-19 recommendation** | <u>N=237</u> | <u>N=196</u> | <u>N=39</u> | <u>N=78</u> | <u>N=38</u> | <u>N=121</u> |
| Before | 214 (90.3%) | 178 (90.8%) | 34.0 (87.2%) | 69.0 (88.5%) | 34.0 (89.5%) | 111 (91.7%) |
| After | 23.0 (9.7%) | 18.0 (9.2%) | 5.00 (12.8%) | 9.00 (11.5%) | 4.00 (10.5%) | 10.0 (8.3%) |

SD = standard deviation
NS-SEC = National Statistics Socioeconomic classification

Demographics of the included respondents are summarised in Table 1. Participant ethnicity was approximately reflective of that of the overall Liverpool City Region with 90.7% listing themselves as “White British”[22-27]. For occupation, 79.7% (n=189/237) of respondents were in more advantaged groups NS-SEC groups 1-4. For household income, 17.9% (n=42/234) had incomes less than or equal to £30,000 and 47.0% (n=110/234) stated their income as between £30,001 and £60,000.

Most of the women who responded to the questionnaire had received the pertussis vaccine (74.7%; n=177/237), and most also took antenatal vitamins (86.9%; n=206/237). The majority of women surveyed were non-smokers (96.2%; n=225/234). The majority of respondents listed themselves as not being in a high-risk group (85.2%; n=202/237) and 81% (n=192/237) did not shield during the COVID-19 pandemic. Health behaviours of the included respondents are summarised in Supplementary file 2, Table S1.

### Attitudes and behaviours towards influenza and influenza vaccine

Most women had received the influenza vaccine in their current pregnancy (73.8%; n=175/237) and 21 (8.9%) had not received the vaccine but intended to do so; 39 women (16.5%) reported that they did not intend to receive the influenza vaccine during their pregnancy (the remaining 2 respondents left this question blank). Of the women who had been vaccinated against influenza or intended to be vaccinated, most (80.6%; n=158/196) had also received the pertussis vaccine (Supplementary file 2, Table S1). One-hundred-five women (46.1%; n=105/228) stated receiving the influenza vaccine during a previous pregnancy, and of these women, 14 (13.3%) said they had experienced side effects.

The attitudes and beliefs of these women towards influenza illness as well as their perceived risks of the virus are summarised in Table 2. Less than half of the women (46.0%; n=109/237) believed that they would get very ill if they got influenza. The majority (70.8%; n=167/236) believed that if they got influenza, their baby could get ill and 64.6% (n=153/237) believed it could hurt their baby. However, just over half of the women perceived themselves as being at risk of getting influenza or that their family/friends were at risk (56.5%; n=134/237 and 57.8%; n=137/237, respectively).

**Table 2.**
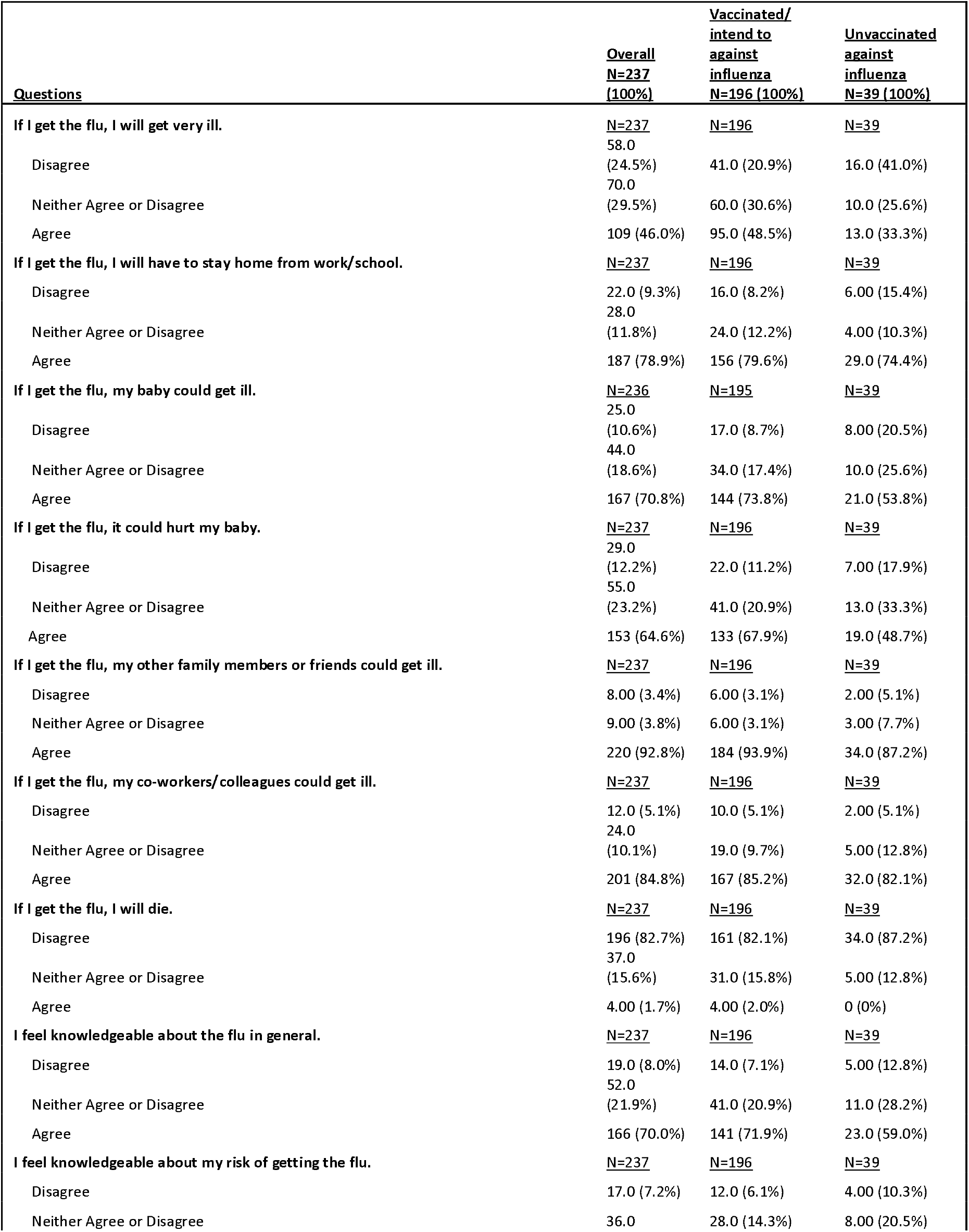
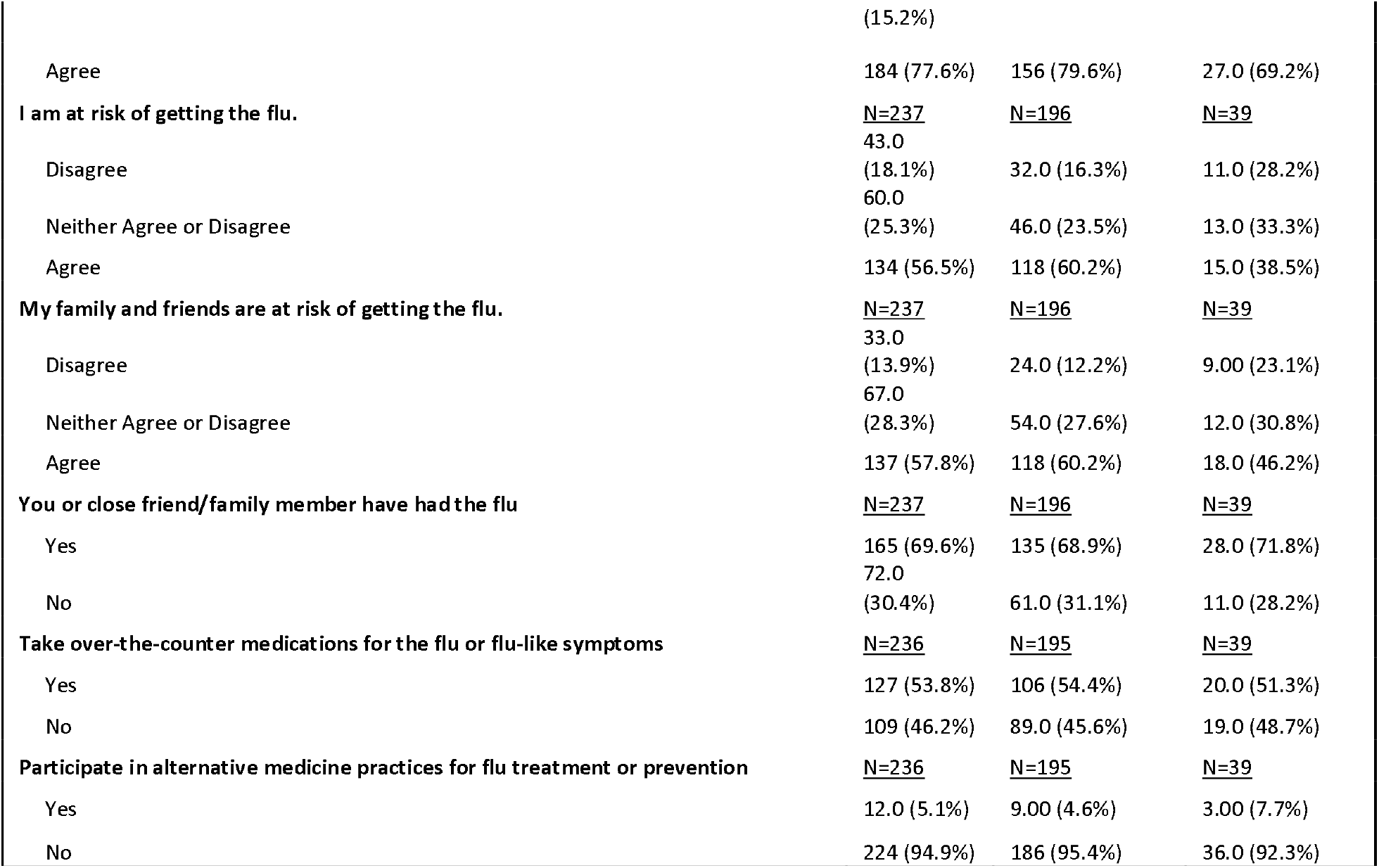
Attitudes and beliefs of pregnant women in the Liverpool City Region, UK towards influenza illness and perceive risks shown in relation to those who are vaccinated/unvaccinated against the virus.

Of the vaccinated women, more than half (60.2%; n=118/196) perceived themselves to be at risk of getting influenza compared to 38.5% (n=15/39) of the unvaccinated women; of vaccinated women, 67.9% (n=133/196) believed that if they got influenza, it could hurt their baby compared to 48.7% (n=19/39) of the unvaccinated women.

The attitudes and beliefs of the pregnant women towards the influenza vaccine are summarised in Table 3. Fifty-three (22.4%) of the 237 women believed they would experience side effects if they receive the influenza vaccine and 99/237 (41.8%) did not believe this. The majority of the women (83.5%; n=198/237) believed that the vaccine would not hurt their baby. In terms of inconvenience or shortages of the vaccine, almost a quarter of the women did not disagree that it was inconvenient for them to receive the vaccine and more than a quarter did not disagree that there was a shortage of the vaccine (24.1%; n=57/237 and 35.0%; n=83/237, respectively) Questions were posed about perceived effectiveness of the influenza vaccine. Of the respondents, more women believed that the influenza vaccine would prevent family members and friends from getting ill and their baby from getting ill than believed it would prevent themselves from getting ill (61.6%; n=146/237, 60.3%; (n=143/237), and 34.2%; n=81/237, respectively). Just over half of the unvaccinated women (51.3%; n=20/39) did not believe that the vaccine is effective at preventing them from getting the virus. More vaccinated women (67.9%; n=133/196) believed that getting the influenza vaccine would help prevent their baby from getting influenza than unvaccinated women (23.1%; n=9/39).

**Table 3.** Attitudes and beliefs of pregnant women in the Liverpool City Region, UK towards the influenza vaccine shown in relation to those who are vaccinated/unvaccinated against the virus.

| <u>Questions</u> | <u>Overall</u><br><u>N=237</u><br><u>(100%)</u> | <u>Vaccinated/</u><br><u>intend to</u><br><u>against</u><br><u>influenza</u><br><u>N=196 (100%)</u> | <u>Unvaccinated</u><br><u>against</u><br><u>influenza</u><br><u>N=39 (100%)</u> |
| --- | --- | --- | --- |
| **If I have the flu vaccine, I will have side effects from it.** | **<u>N=237</u>** | **<u>N=196</u>** | **<u>N=39</u>** |
| Disagree | 99.0 (41.8%) | 89.0 (45.4%) | 9.00 (23.1%) |
| Neither Agree or Disagree | 85.0 (35.9%) | 74.0 (37.8%) | 10.0 (25.6%) |
| Agree | 53.0 (22.4%) | 33.0 (16.8%) | 20.0 (51.3%) |
| **If I have the flu vaccine, I will get ill from it.** | **<u>N=237</u>** | **<u>N=196</u>** | **<u>N=39</u>** |
| Disagree | 177 (74.7%) | 157 (80.1%) | 18.0 (46.2%) |
| Neither Agree or Disagree | 39.0 (16.5%) | 33.0 (16.8%) | 6.00 (15.4%) |
| Agree | 21.0 (8.9%) | 6.00 (3.1%) | 15.0 (38.5%) |
| **If I have the flu vaccine, it could hurt my baby.** | **<u>N=237</u>** | **<u>N=196</u>** | **<u>N=39</u>** |
| Disagree | 198 (83.5%) | 180 (91.8%) | 16.0 (41.0%) |
| Neither Agree or Disagree | 34.0 (14.3%) | 15.0 (7.7%) | 19.0 (48.7%) |
| Agree | 5.00 (2.1%) | 1.00 (0.5%) | 4.00 (10.3%) |
| **If I have the flu vaccine, it will be painful.** | **<u>N=237</u>** | **<u>N=196</u>** | **<u>N=39</u>** |
| Disagree | 177 (74.7%) | 157 (80.1%) | 18.0 (46.2%) |
| Neither Agree or Disagree | 35.0 (14.8%) | 24.0 (12.2%) | 11.0 (28.2%) |
| Agree | 25.0 (10.5%) | 15.0 (7.7%) | 10.0 (25.6%) |
| **If I have the flu vaccine, it will not protect me from getting the flu.** | **<u>N=237</u>** | **<u>N=196</u>** | **<u>N=39</u>** |
| Disagree | 166 (70.0%) | 148 (75.5%) | 17.0 (43.6%) |
| Neither Agree or Disagree | 43.0 (18.1%) | 33.0 (16.8%) | 9.00 (23.1%) |
| Agree | 28.0 (11.8%) | 15.0 (7.7%) | 13.0 (33.3%) |
| **If I have the flu vaccine, it will not protect my baby.** | **<u>N=236</u>** | **<u>N=195</u>** | **<u>N=39</u>** |
| Disagree | 167 (70.8%) | 154 (79.0%) | 11.0 (28.2%) |
| Neither Agree or Disagree | 54.0 (22.9%) | 34.0 (17.4%) | 20.0 (51.3%) |
| Agree | 15.0 (6.4%) | 7.00 (3.6%) | 8.00 (20.5%) |
| **It is inconvenient for me to get the flu vaccine.** | **<u>N=237</u>** | **<u>N=196</u>** | **<u>N=39</u>** |
| Disagree | 180 (75.9%) | 160 (81.6%) | 18.0 (46.2%) |
| Neither Agree or Disagree | 15.0 (6.3%) | 4.00 (2.0%) | 11.0 (28.2%) |
| Agree | 42.0 (17.7%) | 32.0 (16.3%) | 10.0 (25.6%) |
| **There is a shortage of the flu vaccine.** | **<u>N=237</u>** | **<u>N=196</u>** | **<u>N=39</u>** |
| Disagree | 154 (65.0%) | 131 (66.8%) | 22.0 (56.4%) |
| Neither Agree or Disagree | 60.0 (25.3%) | 46.0 (23.5%) | 13.0 (33.3%) |
| Agree | 23.0 (9.7%) | 19.0 (9.7%) | 4.00 (10.3%) |
| **The flu vaccine was recommended to me by my healthcare provider (e.g. doctor, nurse, midwife).** | **<u>N=237</u>** | **<u>N=196</u>** | **<u>N=39</u>** |
| Disagree | 19.0 (8.0%) | 9.00 (4.6%) | 10.0 (25.6%) |
| Neither Agree or Disagree | 21.0 (8.9%) | 12.0 (6.1%) | 8.00 (20.5%) |
| Agree | 197 (83.1%) | 175 (89.3%) | 21.0 (53.8%) |
| **If I have the flu vaccine, I will not get ill with the flu.** | **<u>N=237</u>** | **<u>N=196</u>** | **<u>N=39</u>** |
| Disagree | 87.0 (36.7%) | 67.0 (34.2%) | 20.0 (51.3%) |
| Neither Agree or Disagree | 69.0 (29.1%) | 59.0 (30.1%) | 10.0 (25.6%) |
| Agree | 81.0 (34.2%) | 70.0 (35.7%) | 9.00 (23.1%) |
| **If I have the flu vaccine, I will help prevent my baby from getting the flu.** | **<u>N=237</u>** | **<u>N=196</u>** | **<u>N=39</u>** |
| Disagree | 42.0 (17.7%) | 26.0 (13.3%) | 15.0 (38.5%) |
| Neither Agree or Disagree | 52.0 (21.9%) | 37.0 (18.9%) | 15.0 (38.5%) |
| Agree | 143 (60.3%) | 133 (67.9%) | 9.00 (23.1%) |
| **If I have the flu vaccine, I will help prevent my family/friends from getting ill with the flu.** | **<u>N=237</u>** | **<u>N=196</u>** | **<u>N=39</u>** |
| Disagree | 48.0 (20.3%) | 34.0 (17.3%) | 14.0 (35.9%) |
| Neither Agree or Disagree | 43.0 (18.1%) | 31.0 (15.8%) | 12.0 (30.8%) |
| Agree | 146 (61.6%) | 131 (66.8%) | 13.0 (33.3%) |
| **Received flu vaccine during previous pregnancy** | **<u>N=228</u>** | **<u>N=187</u>** | **<u>N=39</u>** |
| Yes | 105 (46.1%) | 97.0 (51.9%) | 7.00 (17.9%) |
| No | 123 (53.9%) | 90.0 (48.1%) | 32.0 (82.1%) |
| **If received flu vaccine in previous pregnancy, were there side effects?** | **<u>N=105</u>** | **<u>N=97</u>** | **<u>N=7</u>** |
| Yes | 14.0 (13.3%) | 13.0 (13.4%) | 1.00 (14.3%) |
| No | 91.0 (86.7%) | 84.0 (86.6%) | 6.00 (85.7%) |

The means by which the pregnant women were offered the influenza vaccine is shown in Supplementary file 2, Table S2. The vast majority of pregnant women (89.9%; n=213/237) reported being offered the influenza vaccine and 44.7% (n=106/237) had it offered by their general practitioner and 46% (n=109/237) by community services/midwife. More than half (58.2%; n=138/237) were offered the vaccine in a face-to-face setting.

### Attitudes and behaviours towards vaccines in general

The attitudes and beliefs of the pregnant women towards vaccines in general are shown in Supplementary file 2, Table S3. Most of the respondents across all categories believed vaccines to prevent disease and many more of those who were vaccinated/intended to vaccinate against influenza believed vaccines to be safe than those who were unvaccinated (83.7%; n=164/196 and 56.4%; n=22/39, respectively). A third (33.3%; n=13/39) of those who were unvaccinated against influenza intended to vaccinate their child against influenza when they are old enough, however most of the women across all categories intended to vaccinate their baby when they are born with all vaccines offered.

The self-reported likelihood of the pregnant women in this study accepting a vaccine varied by the type of healthcare professionals making the recommendation (Supplementary file 2, Table S4). For the healthcare professionals listed, 85.2% (n=201/236) would accept it from a doctor, 68.0% (n=157/231) from a pharmacist, 77.0% (n=181/235) from a nurse, 84.4% (n=200/237) from a midwife, and 70.1% (n=164/234) from a health visitor.

### Attitudes and behaviours towards COVID-19 and COVID-19 vaccine

Of the 237 respondents, 34.2% (n=81) believed they would get very ill if they got COVID-19 and 20.3% (n=48) disagreed (Table 4). Almost all of the participants believed they would have to isolate if they became ill with COVID-19 (99.6%; n=236/237) and all of them believed that if they became ill, their family members and friends with whom they came into contact would have to quarantine. Over three-quarters of the women (78.5%; n=186/237) believed that if they became ill with COVID-19, their baby could get ill.

**Table 4.**
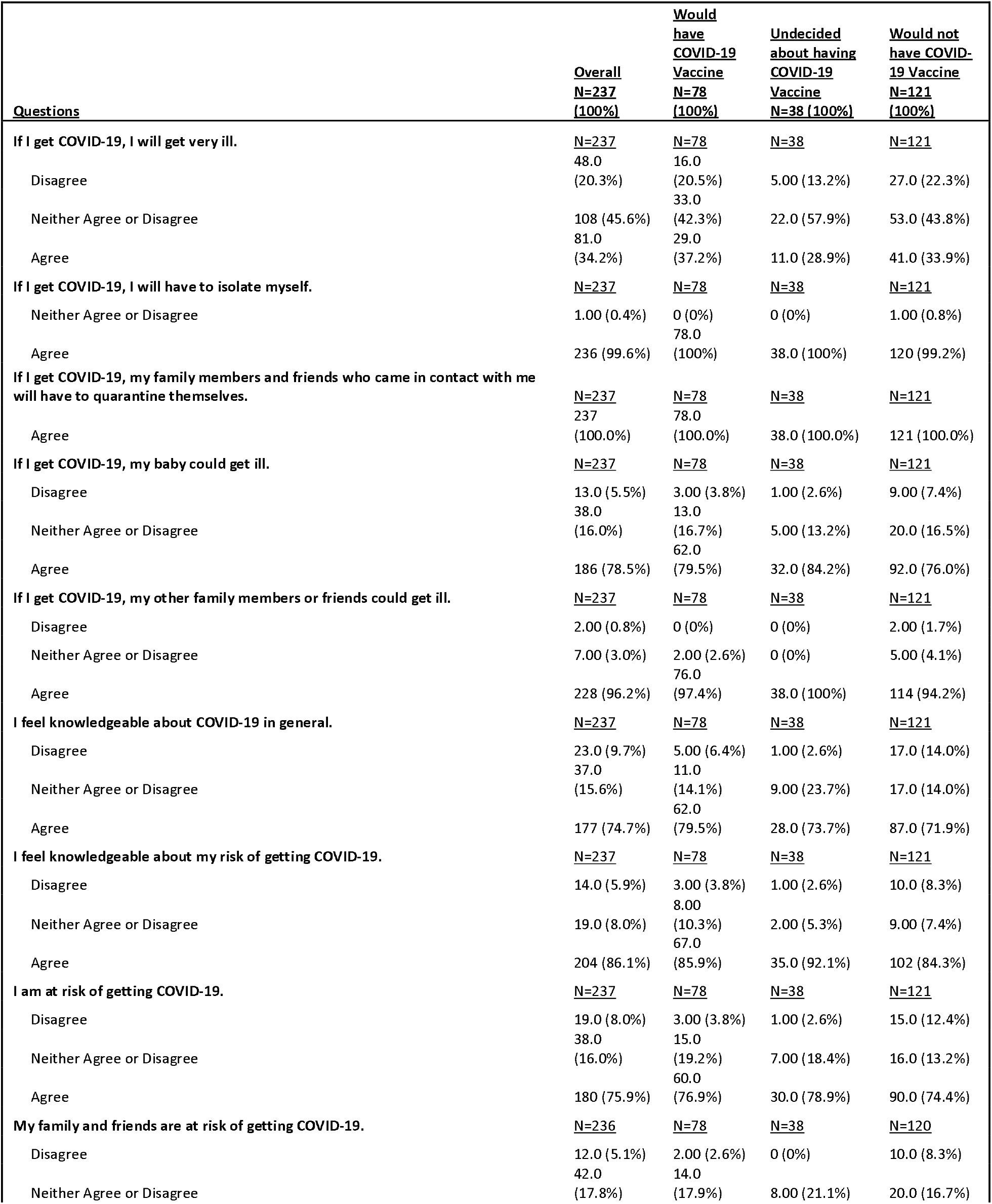
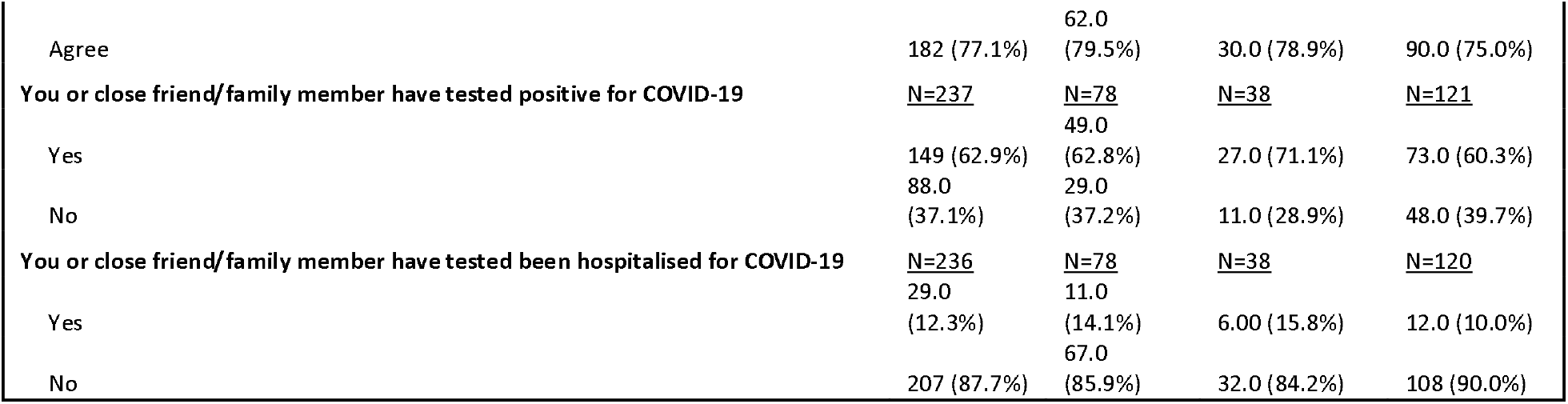
Attitudes and beliefs of pregnant women in the Liverpool City Region, UK towards COVID-19 illness and perceived risks shown in relation to possible COVID-19 vaccine acceptance.

Most of the women stated that they believed themselves to be knowledgeable about the COVID-19 disease and their risks (74.7%; n=177/237 and 86.1%; n=204/237, respectively); most also perceived themselves to be at risk of getting ill with the disease (75.9%; n=180/237). Many of the respondents (62.9%; n=149/237) had previously tested positive for COVID-19 or had a close friend or family member test positive for COVID-19 (Table 4).

The attitudes and beliefs of the pregnant women participating in this study towards the COVID-19 vaccine are summarised in Table 5. Most of the responses (90.3%; Table 1) to this questionnaire were received before the approval of pregnant women to receive the COVID-19 vaccine on the 16^th^ of April, 2021. Of the respondents, fewer of them were willing to receive a COVID-19 vaccine if it were available to them than those who were willing (32.9%; n=78/237 and 51.1%; n=121/237, respectively). However, the vast majority reported they would be willing to receive the vaccine if they were not pregnant (80.2%; n=190/237). Of those who would be willing to accept the COVID-19 vaccine, 87.2% (n=68/78) had received the pertussis vaccine, which is slightly higher than those who had been vaccinated/intended to be vaccinated against influenza (Supplementary file 2, Table S1). Unlike with the influenza vaccine, more women believed that the COVID-19 vaccine would protect themselves than believed it would protect their baby, other family members, or friends (65.0%; n=154/237 and 54.9%; n=130/237, respectively). Eighty-six of the 237 women (36.3%) said they would vaccinate their baby against COVID-19 as soon as possible after they were born, while 35% said (n=83/237) they would not. Most of the women (68.6%; n=162/236) said they would get the COVID-19 vaccine every year if it were a seasonal vaccine. When comparing willingness to have the COVID-19 vaccine vs. if participants had/intend to have the influenza vaccine, most responses fell under unwillingness to receive the COVID-19 vaccine or the neither willing nor unwilling regardless of whether the women had received or intended to receive the influenza vaccine (47.4%; n=93/196 for those who had/intend to have the influenza vaccine and 71.8%;n=28/39 for those who had not).

**Table 5.**
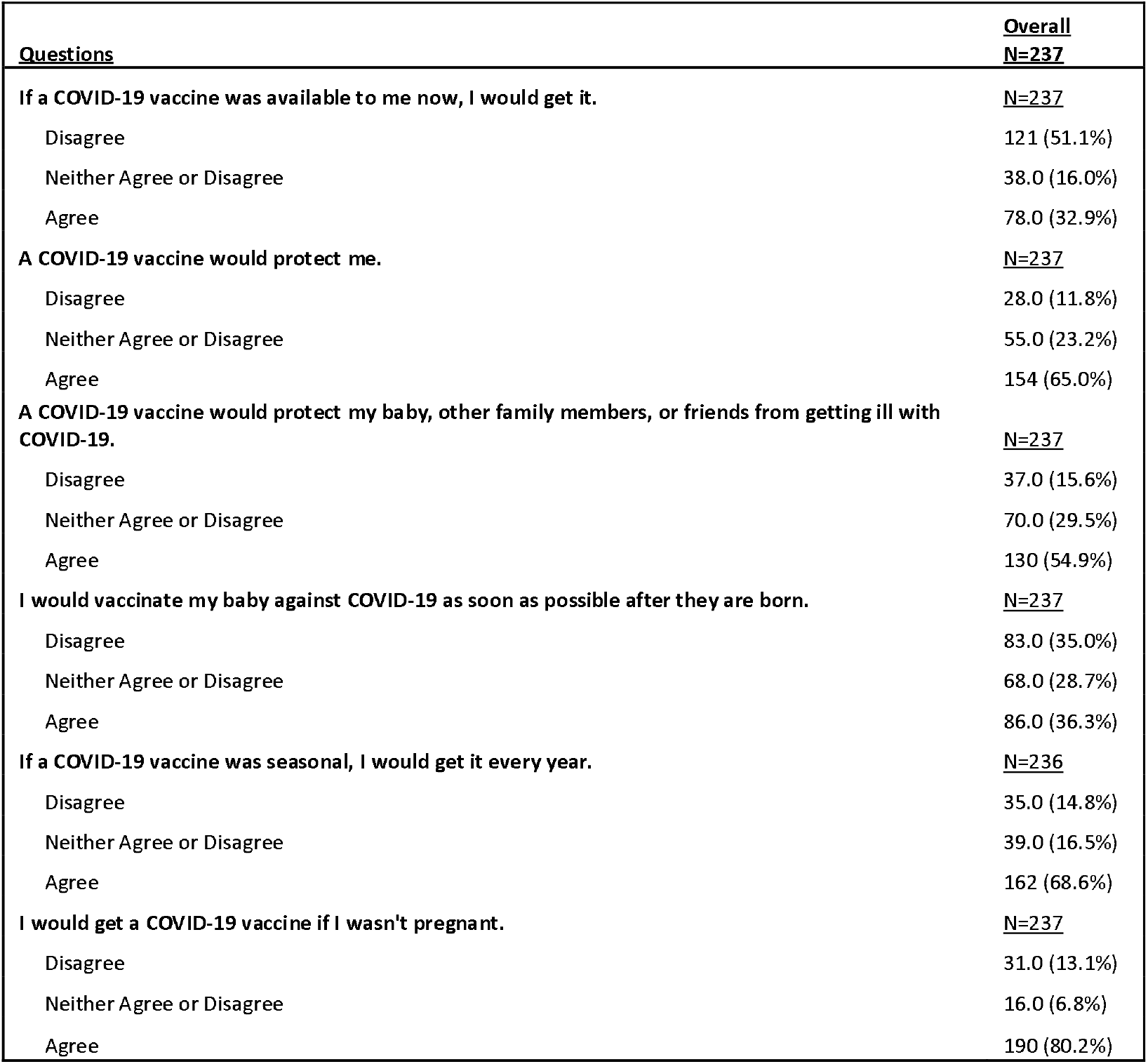
Attitudes and beliefs of pregnant women in the Liverpool City Region, UK towards the COVID-19 vaccine.

The likelihood of the pregnant women accepting the COVID-19 vaccine if recommended to them by different healthcare professionals is shown in Supplementary file 2, Table S5. Most women were willing to accept the COVID-19 vaccine if recommended to them by a doctor, nurse, or midwife (78.0%; n=184/236, 64.6%; n=153/230, and 73.5%; n=172/234, respectively) while less were willing to accept it if recommended by a pharmacist or health visitor (57.5%; n=131/228 and 59.3%; n=137/231, respectively).

## Discussion

The findings of this cross-sectional survey indicate that the majority of respondents had received the influenza vaccine and most perceived the influenza virus as causing more harm to their baby rather than to themselves. For the attitudes about COVID-19 and the COVID-19 vaccine, only about a third of the women believed they would get very ill from the disease. This shows the importance of providing information about the effects of viruses like SARS-CoV2 and influenza on pregnant women specifically. Unlike with the influenza illness questions, there was less of a divergence in responses to COVID-19 questions; most of the women felt knowledgeable about the disease and its risks and perceived themselves and their friends and family to be at risk. This is most likely due to the extensive media coverage of COVID-19. Additionally, many of the women stated they would receive the COVID-19 vaccine if they were not pregnant; this may be due to the women not having accessible information regarding the safety of the vaccine during pregnancy.

Similar to our finding, a previous study the UK found that the two main reasons behind hesitancy in pregnant women towards the influenza vaccine were that pregnant women were more likely to implement healthy behaviours (such as not smoking) if they benefitted the baby rather than themselves and that there is a misconception that maternal morbidity and mortality from influenza infection is low[28].

In general, the influenza vaccine uptake for Liverpool City Region is low (<50%), but for the pregnant women in this study, there was relatively high uptake (82.7% received/intended to receive). Therefore, we have likely accessed a subgroup of pregnant women more inclined towards maternal vaccination, providing information on what drives uptake.

In this group of pregnant women, more than half of them did not agree that they would get very ill from influenza and while most believed that getting influenza could hurt their baby, there was still a group of women that did not fall into that “Agree” category. It is important to understand why some women didn’t agree with the statement. It could be related to the information available regarding vaccination or the way that the information is provided about the virus and its risks to them and their baby.

The majority of the women in this study reported feeling knowledgeable about influenza and their risk of getting influenza, however, only just over half felt that they are at risk of getting influenza; understanding the reasons for the divergence of the responses to these two questions is necessary. Influenza is an under-rated health problem in pregnant women, and there is a lack of belief that the vaccine will protect them from influenza infection[29]. A cross-sectional survey that was done in Saudi Arabia found that when pregnant women had a poor knowledge of influenza and the safety of the vaccine during pregnancy, they were significantly less likely to accept it[30]. Another study from Australia showed that one of the main barriers for not receiving the influenza vaccine during their pregnancy was lack of recommendation from medical staff[31]. A Spanish study stressed the influence of medical professionals’ recommendations of influenza vaccination on pregnant women’s acceptance of the vaccine[32]. The higher morbidity and mortality effects can be lessened with the aid of education of pregnant women about influenza vaccination[29]. These studies stress the importance of medical staff recommendations and the quality of influenza vaccination information provided to pregnant women on acceptance of the vaccine. Interestingly, most of the women in our study had the vaccine recommended to them by their healthcare provider, which may partly explain why the proportion of women vaccinated was much higher in our study compared to the population average.

As the questionnaire was conducted during the COVID-19 pandemic and a national lockdown in the UK, it can provide insight into the potential effects of the pandemic on attitudes towards influenza vaccination in pregnancy and vaccines in general. Survey distribution occurred mainly before the COVID-19 vaccine was approved for use in pregnant women. Since more of the women reported that they would receive the vaccine if they were not pregnant that those who would receive it during their pregnancy, it is likely that some of the hesitancy towards acceptance of the COVID-19 vaccine during pregnancy is due to the limited amount of studies in the area; it also seems that this hesitancy in our study group is more related to the COVID-19 vaccine specifically than to all maternal vaccinations. Indeed, a nationwide cross-sectional survey in Qatar found that 25% of perinatal women were hesitant about receiving a COVID-19 vaccine citing infection risks and safety concerns [33]. A web survey conducted multi-nationally (including the UK) found that of the pregnant and breastfeeding women sampled, hesitancy towards the COVID-19 vaccine was found in 40-50%[34]. More rapid cycle evaluation will need to be conducted on COVID-19 vaccine safety and effectiveness in pregnancy and disseminated efficiently as the safety of the baby is the priority for this group of women. These studies stress the importance of providing timely and accessible information to pregnant women about vaccines during pregnancy. Our study also shows the importance of who provides this information to pregnant women, with doctors and midwives most influential.

## Strengths and Limitations

Limited scientific research has been conducted with SARS-CoV2 virus and the COVID-19 disease in pregnant women and clinical trials of the vaccine have yet to be reported in pregnant women. Also, understanding the attitudes of pregnant women toward influenza vaccination, the reasoning for hesitancy, and the importance of medical staff recommendations can assist in developing messages that provide information on the safety and effectiveness of the vaccine. The study raises some contradictions in people’s beliefs which is worthy of further investigation.

Additionally, some of the questions posed limited the responses we could receive. For example, a question was asked regarding the number of children a participant currently had and not the number of pregnancies, therefore limiting the possibility of miscarriage. Future surveys will ask questions about the number of pregnancies as well as the number of children to account for this limitation.

The small sample size, the self-reflected nature of the sample, and the high uptake of the influenza vaccine in this sample of pregnant women raise questions about the extent to which the sample is representative of pregnant women living in the geographical region of Liverpool. Given that the influenza vaccine status was self-reported and, because of the anonymous nature of the survey, could not be confirmed through health records, it is possible that the reported uptake of the influenza vaccine overestimated the number of women immunised or intending to be immunised. Due to the COVID-19 pandemic, the survey was largely distributed through social media links so we were unable to access women who did not use the social media sites we selected. Future surveys could use NHS sites (with appropriate approvals) to access the pregnant woman population to achieve a more representative population. Finally, due to the small sample size, subgroup analyses of ethnically diverse groups and less advantaged groups were not possible; this small sample size also made statistical analyses inappropriate.

## Conclusions

Most of the women in this study had received the influenza vaccine during their pregnancy. However, the concerns they had were more related to the safety of their baby rather than themselves. Correct and accessible information on the risks of influenza illness as well as vaccine recommendation, especially from doctors and midwives, plays a huge part in perceptions of vaccine effectiveness and safety in pregnant women. Vaccine hesitancy towards COVID-19 vaccines in this group of pregnant women from Liverpool, UK seem to be as a result of not knowing the risks to them or their child of both COVID-19 illness and potential side effects/adverse events from COVID-19 vaccination. This is likely directly related to the paucity of scientific studies in the area. Future surveys will ask for reasons behind potential hesitancy in order to gain a better understanding. Further studies will need to be conducted, perhaps focusing on less advantaged, hard to reach groups, to further understand the perceptions of influenza and COVID-19 vaccine safety in pregnancy. The next steps in this study will be to conduct focus groups and interviews to gain a deeper understanding of attitudes towards and enablers of influenza and COVID-19 vaccination in pregnancy.

## Supporting information

Supplementary file 1. Questionnaire.

Supplementary file 2. Supplementary images and tables.

## Data Availability

The data in this study was collected via a survey from members of the public. As the study participants did not explicitly consent to their data being made available in a public repository, we have been advised by the University of Liverpool Research Data Management Team that it is not appropriate to make the data available in this format. However, datasets used and/or analysed during the current study may be available from the corresponding author on reasonable request.

## Acknowledgements

The authors would like to thank the respondents of the online questionnaire and the patient and public engagement panel, as well as the multiple groups through social media and the Liverpool Babies Patient and Public Involvement and Engagement Group for their aid in distributing the questionnaire through their various networks.

## Patient Consent for Publication

Not required.

## Ethics Approval

The project received ethical approval from the University of Liverpool Health and Life Sciences Research Ethics Committee (Human participants, tissues and databases) (approval number: 7865). The online survey was anonymous and valid informed consent was gained from study participants prior to completion of the questionnaire; recoded using clear affirmative action by use of a consent checkbox. The ethics committee approved this method of valid informed consent.

## Author Contributions

DH, SK, NF, EP conceptualised and designed the study. DH, NF, EP were responsible for supervision. SK and DH were responsible for survey design, investigation, and verifying the data. SK was responsible for visualisation, formal analysis, and writing the original draft. All authors reviewed and edited the manuscript. The authors read and approved the final manuscript.

