## Supplementary file 1. Questionnaire. for "Pregnant women’s attitudes and behaviours towards antenatal vaccination against Influenza and COVID-19 in the Liverpool City Region, United Kingdom: cross-sectional survey"

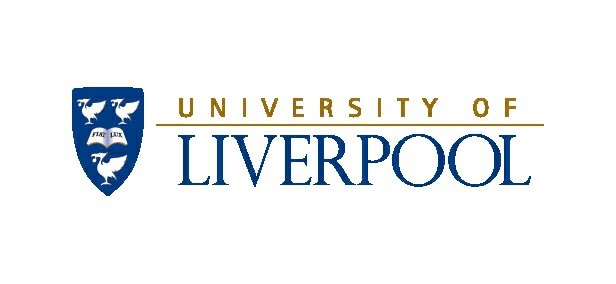

Flu and COVID-19 Attitudes and Behaviours

Page 1: Flu and COVID-19 Attitudes and Behaviours

This questionnaire is	about	the flu	(Influenza) and the flu vaccine which is offered during pregnancy and to children aged 2	and older. It	is also	about COVID-19	(SARS-CoV-2).

The questionnaire is	part of	a research project that aims to look at knowledge	and attitudes	about the flu	and its vaccine as well as about COVID-19. Th	information gathered from the questionnaire will aid in developing an	informative message about influenza and its vaccine in an effort to increase flu vaccine uptake. It will also provide information on the effect of COVID-19 and its	pandemic on flu vaccine acceptance.

All of the responses to this questionnaire	are confidential and anonymous (no	information will be gathered that could identify	you). Participation in this questionnaire is completely voluntary and you may choose to leave the survey at any	time if you feel you do not wish to complete it. Since the questionnaire is	confidential, once submitted, no responses can be withdrawn as it will be	impossible to find which responses relate to you. Data collected from this	questionnaire will be retained for 10 years on the secure University network	and then deleted.

When the questionnaire is completed, you will be provided with contact details for	the study if you have any questions.

If you have already completed this questionnaire, please do not complete it again. **Additionally, please only complete this questionnaire if you are pregnant and if you live in the Liverpool City Region (Liverpool, Knowsley, Sefton, St Helens, Wirral, Halton).**

This questionnaire is voluntary and as such, if a question makes you uncomfortable,	you may choose to leave it blank. However, we ask that you answer as many of the	questions as you can.

This questionnaire will take approximately 10 minutes to complete.

If you have any questions, please	contact:

Samantha Kilada

Department of Clinical Infection, Microbiology and Immunology,

University of Liverpool

Ronald Ross Building, 8 West Derby Street

Liverpool, L69 7BE

1. **Please tick the box to indicate your consent to the information above.**	

*Required*

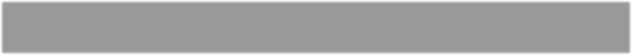

I consent.

Page 2:

Background Information

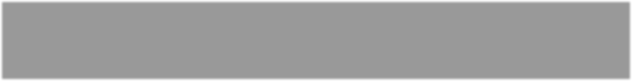

Please enter a	number.

*2.*

How old are you (in years)?

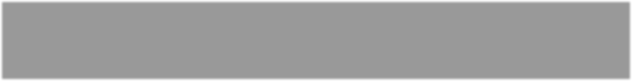

Please enter a	number.

*3.*

Approximately how many weeks pregnant are you?

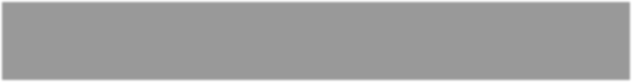

This is my first

1

2

3

4

5+

*4.*

How many children do you currently have?

*5.*	Are you a member of a high-risk health group (e.g. asthma, liver disease, diabetes, etc.)?

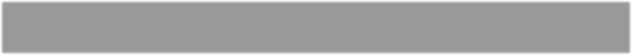

Yes

No

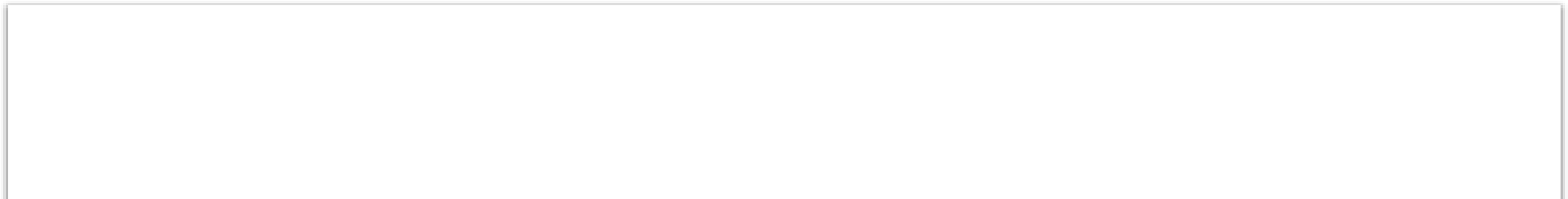

Below GCSE

GCSE or similar

NVQ or similar

A-level or similar

Undergraduate

Post-graduate

*6.*

What is your highest education level?

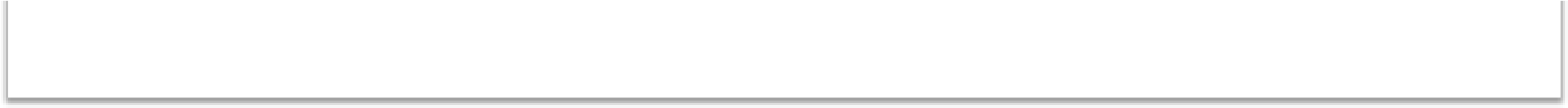

Other

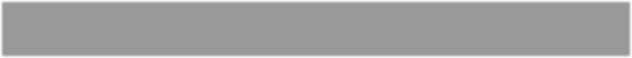

*6*

*.a.*

If you selected Other, please specify:

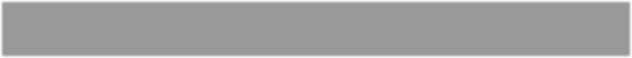

*7.*

What is your current occupation?

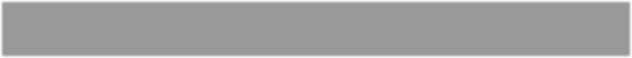

*8.*

What is the first part of your postcode (e.g. “L17”)

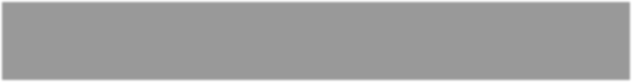

<

£

10,000

£10,001-20,000

£20,001-30,000

£30,001-45,000

£45,001-60,000

>

£

60,000

*9.*

What is your estimated family income?

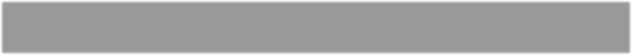

Yes

No

*10.*

Do you own a smartphone?

*11.* Do you access your GP online for any of these services? (check all that apply)

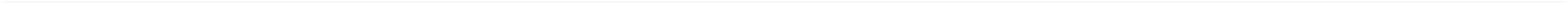

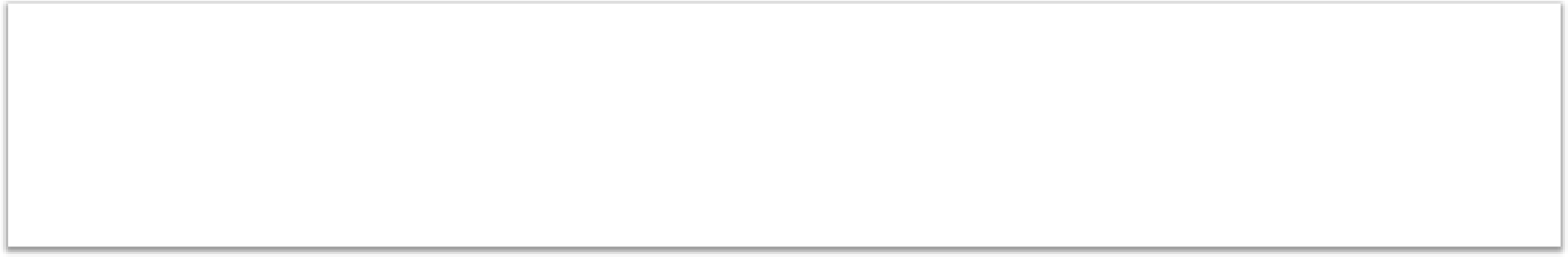

Book	appointments

Request	prescriptions

Access	medical

records

Other

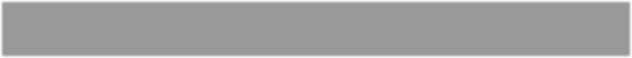

*11*

*.a.*

If you selected Other, please specify:

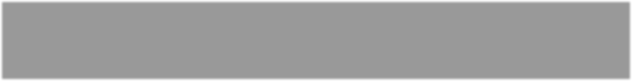

Please enter a number.

*12.*

How many people live in your household (including yourself)?

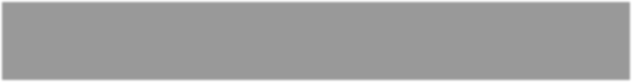

Alone

Partner

Child(ren)

Parent(s)

Other	family

Roommate(s)

*13.*

Who do you live with? (check all that apply)

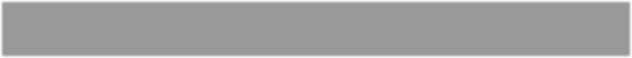

*14.*

What is your ethnicity?

Page 3: Health Behaviours

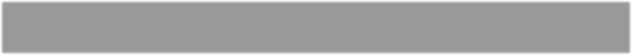

Yes

No

*15.*

Do you currently smoke?

16. Prior to pregnancy, how many days (on	average) per week did you	do at	least 30 minutes of exercise?

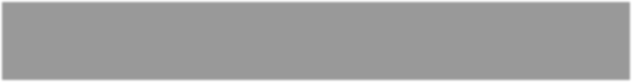

0

1-2

3-4

5-7

17. Currently, how many days (on average) per week do you do at least 30	minutes of exercise?

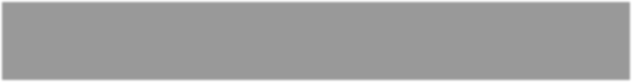

0

1-2

3-4

5-7

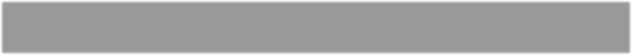

Yes

No

*18.*

Do you take antenatal vitamins?

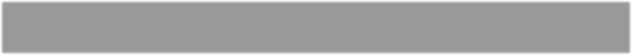

Yes

No

*19.*

Do you take folic acid?

Page 4: Flu Illness

**Please	read these statements and then tick only one box for each statement.**

20. **This question is in	relation to your beliefs about flu	illness. Please rate the extent to which you agree with the following statements.**

Please	don't	select	more	than	1	answer(s)	per	row.

|  | Strongly Disagree | Disagree | Neither  Agree or  Disagree | Agree | Strongly  Agree |
| --- | --- | --- | --- | --- | --- |
| If I get the flu, I will get very ill. |  |  |  |  |  |
| If I get the flu, I will have to stay home from work/school. |  |  |  |  |  |
| If I get the flu, my baby could get ill. |  |  |  |  |  |
| If I get the flu, it could hurt my baby. |  |  |  |  |  |
| If I get the flu, my other family members or friends could get ill. |  |  |  |  |  |
| If I get the flu, my coworkers/colleagues could get ill. |  |  |  |  |  |
| If I get the flu, I will die. |  |  |  |  |  |

21. .

Please don't select more than 1	answer(s) per row.

|  | Strongly Disagree | Disagree | Neither  Agree or  Disagree | Agree | Strongly  Agree |
| --- | --- | --- | --- | --- | --- |
| I feel knowledgeable  about the flu in general. |  |  |  |  |  |
| I feel knowledgeable about my risk of getting the flu. |  |  |  |  |  |
| I am at risk of getting the flu. |  |  |  |  |  |
| My family and friends are at risk of getting the flu. |  |  |  |  |  |

Page 5:

Flu Illness and Vaccine

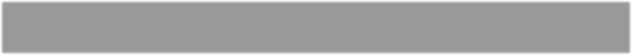

Yes

No

*22.*

Have you been offered the flu this year?

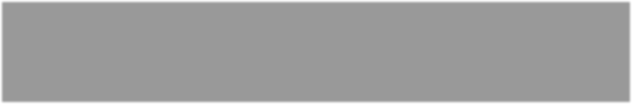

GP

Women's	Hospital

Community

services/midwife

Health	visitor

Pharmacist

Other

*22*

*.a.*

If yes, who offered it to you? (check all that apply)

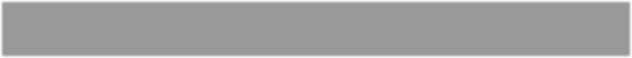

*.a.i.*

*22*

If you selected Other, please specify:

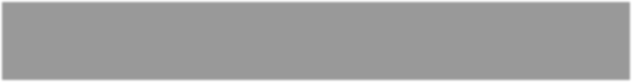

Letter

Text	message

Face-to-face

Other

*22*

*.b.*

How was it offered to you? (check all that apply)

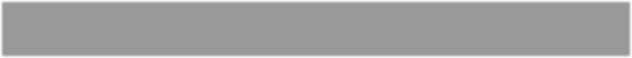

*22*

*.b.i.*

If you selected Other, please specify:

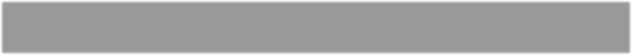

Yes

No

*23.*

Have you had the flu vaccine during this pregnancy??

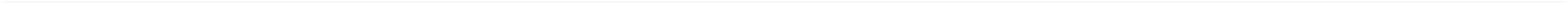

*23*

*.a.*

If you have not had the flu vaccine, do you intend to?

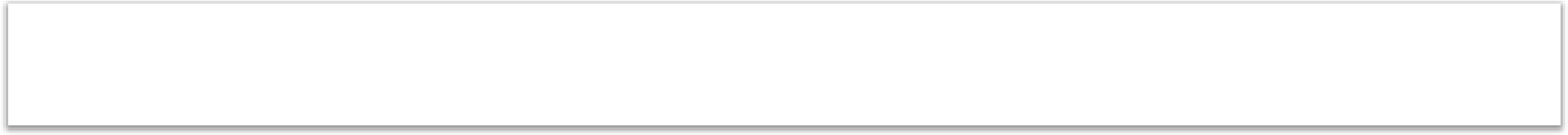

Yes

No

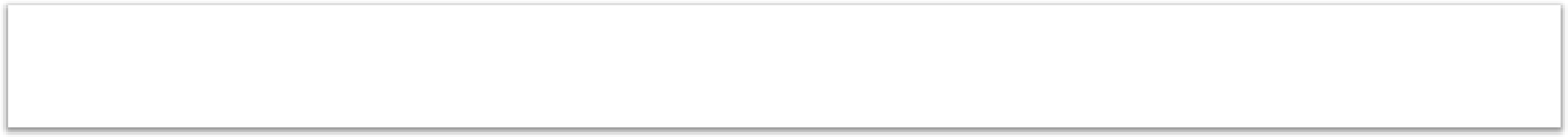

Yes

No

*24.*

Have you had the flu vaccine during a previous pregnancy?

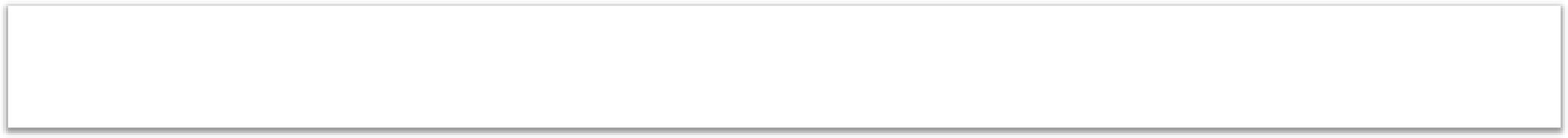

Yes

No

*24*

*.a.*

If you did, did you experience any side effects afterwards?

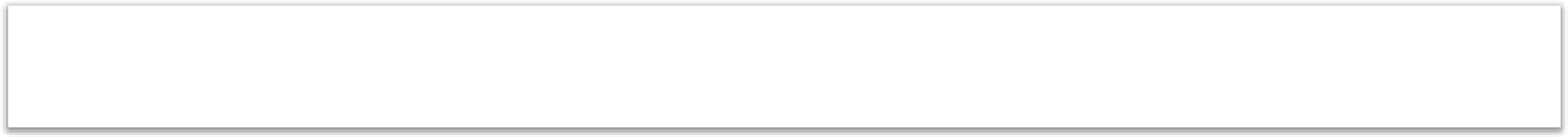

Yes

No

*25.*

Do you discuss vaccination with your partner or another family member?

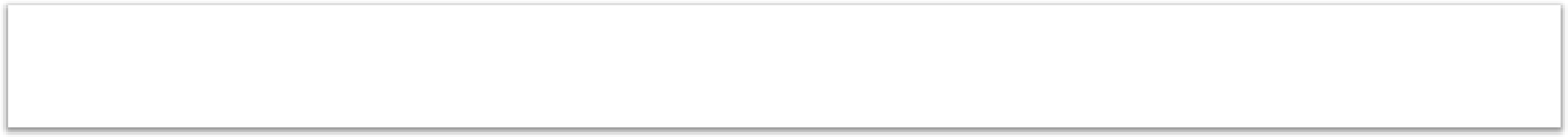

Yes

No

*26.*

Have you or a close friend or family member of yours ever had the flu?

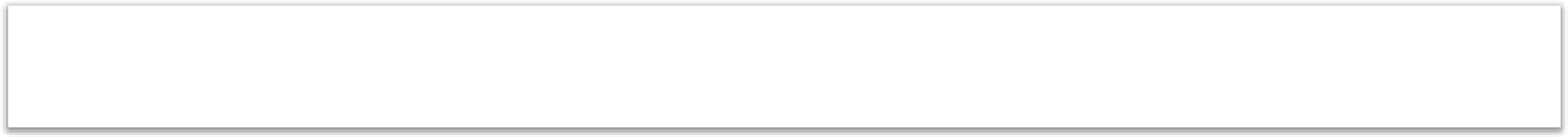

Yes

No

*27.*

Do you take any over-the-counter medications for flu or flu-like symptoms?

*28.*	Do you participate in any alternative medicine practices for flu treatment or prevention?

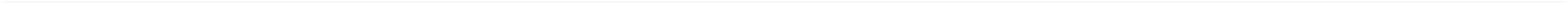

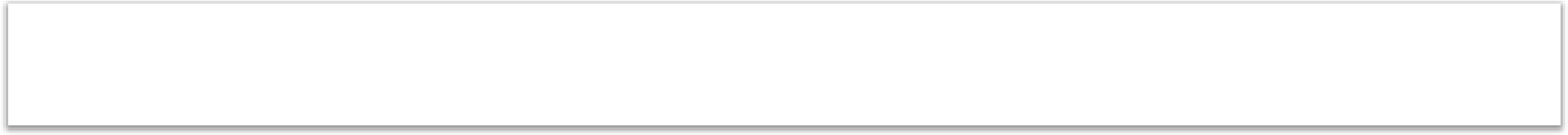

Yes

No

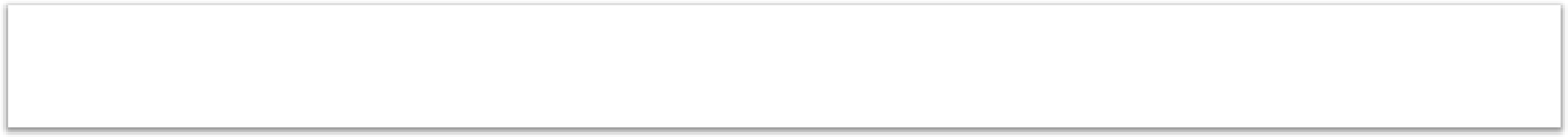

Yes

No

*29.*

Have you had the pertussis (whooping cough) vaccine?

Page 6: Feelings about the Flu Vaccine

**Please read these statements and then tick only one box for each statement.**

30. **This question is in relation to your beliefs about flu vaccine. In the literature, people are reported to hold beliefs that may not be supported by scientific evidence. We have used these ideas to create these questions. Please rate the extent to which you agree with the following statements.**

Please don't select more than 1 answer(s) per row.

|  | Strongly Disagree | Disagree | Neither  Agree or  Disagree | Agree | Strongly  Agree |
| --- | --- | --- | --- | --- | --- |
| If I have the flu vaccine, I will have side effects from it. |  |  |  |  |  |
| If I have the flu vaccine, I will get ill from it. |  |  |  |  |  |
| If I have the flu vaccine, it could hurt my baby. |  |  |  |  |  |
| If I have the flu vaccine, it will be painful. |  |  |  |  |  |
| If I have the flu vaccine, it will not protect me from getting the flu. |  |  |  |  |  |
| If I have the flu vaccine, it will not protect my baby. |  |  |  |  |  |

*30.a.*	.

Please	don't	select	more	than	1	answer(s)	per	row.

|  | Strongly Disagree | Disagree | Neither  Agree or  Disagree | Agree | Strongly  Agree |
| --- | --- | --- | --- | --- | --- |
| It is inconvenient for me to get the flu vaccine. |  |  |  |  |  |
| There is a shortage of the flu vaccine. |  |  |  |  |  |
| The flu vaccine was recommended to me by my healthcare provider (e.g. doctor, nurse, midwife). |  |  |  |  |  |

31. **This question is in relation to your beliefs about flu illness. Please rate the extent to which you agree with the following statements.**

Please don't select more than 1 answer(s) per row.

|  | Strongly Disagree | Disagree | Neither  Agree or  Disagree | Agree | Strongly  Agree |
| --- | --- | --- | --- | --- | --- |
| If I have the flu vaccine, I will not get ill with the flu. |  |  |  |  |  |
| If I have the flu vaccine, I will help prevent my baby from getting the flu. |  |  |  |  |  |
| If I have the flu vaccine, I will help prevent my family/friends from getting ill with the flu. |  |  |  |  |  |

Page 7: Vaccines

**Please read these statements and then tick only one box for each statement.**

32. .

Please don't select more than 1 answer(s) per row.

|  | Strongly Disagree |  | Disagree | Neither  Agree or  Disagree | Agree | Strongly  Agree |
| --- | --- | --- | --- | --- | --- | --- |
| Vaccines prevent disease. |  |  |  |  |  |  |
| Vaccines are safe. |  |  |  |  |  |  |
| I intend to vaccinate my child with the flu vaccine when they are old enough. |  |  |  |  |  |  |
| I intend to vaccinate my baby when they are born with all vaccines offered. |  |  |  |  |  |  |
| I am more likely to have a vaccine if my family members or friends have had it. |  |  |  |  |  |  |

33. **I am more likely to have a vaccine if it is recommended by a:**

Please don't select more than 1 answer(s) per row.

|  | Strongly Disagree | Disagree | Neither  Agree or  Disagree | Agree | Strongly  Agree |
| --- | --- | --- | --- | --- | --- |
| - doctor |  |  |  |  |  |
| - pharmacist |  |  |  |  |  |
| - nurse |  |  |  |  |  |
| - midwife |  |  |  |  |  |
| - health visitor |  |  |  |  |  |
| - family member or friend |  |  |  |  |  |

Page 8: COVID-19

34. Have you or a close friend or family member of yours ever tested positive for COVID-19?

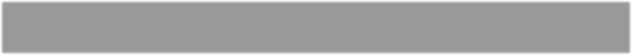

Yes

No

35. Have you or a close friend or family member of yours been hospitalized for	COVID-19?

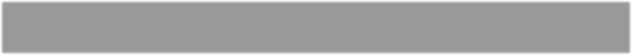

Yes

No

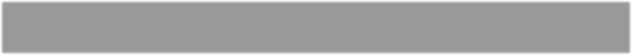

Yes

No

*36.*

Were you shielding during the COVID-19 pandemic?

Page 9: COVID-19 Illness

**Please read these statements and then tick only one box for each statement.**

37. **This question is in relation to your beliefs about COVID-19 illness. Please rate the extent to which you agree with the following statements.**

Please don't select more than 1 answer(s) per row.

|  | Strongly Disagree | Disagree | Neither  Agree or  Disagree | Agree | Strongly  Agree |
| --- | --- | --- | --- | --- | --- |
| If I get COVID-19, I will get very ill. |  |  |  |  |  |
| If I get COVID-19, I will have to isolate myself. |  |  |  |  |  |
| If I get COVID-19, my family members and friends who came in contact with me will have to quarantine themselves. |  |  |  |  |  |
| If I get COVID-19, my baby could get ill. |  |  |  |  |  |
| If I get COVID-19, my other family members or friends could get ill. |  |  |  |  |  |

38. .

Please don't select more than 1 answer(s) per row.

|  | Strongly Disagree | Disagree | Neither  Agree or  Disagree | Agree | Strongly  Agree |
| --- | --- | --- | --- | --- | --- |
| I feel knowledgeable about COVID-19 in general. |  |  |  |  |  |
| I feel knowledgeable about my risk of getting COVID-19. |  |  |  |  |  |
| I am at risk of getting COVID-19. |  |  |  |  |  |
| My family and friends are at risk of getting COVID-19 |  |  |  |  |  |

Page 10: COVID-19 Illness and Vaccine

**Please read these statements and then tick only one box for	each statement.**

39. .

Please don't select more than 1 answer(s) per row.

|  | Strongly Disagree | Disagree | Neither  Agree or  Disagree | Agree | Strongly  Agree |
| --- | --- | --- | --- | --- | --- |
| If a COVID-19 vaccine was available to me now, I would get it. |  |  |  |  |  |
| A COVID-19 vaccine would protect me. |  |  |  |  |  |
| A COVID-19 vaccine would protect my baby, other family members, or friends from getting ill with  COVID-19. |  |  |  |  |  |
| I would vaccinate my baby against COVID-19 as soon as possible after they are born. |  |  |  |  |  |
| If a COVID-19 vaccine was seasonal, I would get it every year. |  |  |  |  |  |
| I would get a COVID-19 vaccine if I wasn't pregnant. |  |  |  |  |  |

40. **I am more likely to have a COVID-19 vaccine if it is recommended to me by a:**

Please don't select more than 1 answer(s) per row.

|  | Strongly Disagree | Disagree | Neither  Agree or  Disagree | Agree | Strongly  Agree |
| --- | --- | --- | --- | --- | --- |
| - doctor. |  |  |  |  |  |
| - pharmacist. |  |  |  |  |  |
| - nurse. |  |  |  |  |  |
| - midwife. |  |  |  |  |  |
| - health visitor. |  |  |  |  |  |
| - family member or friend |  |  |  |  |  |

Page 11

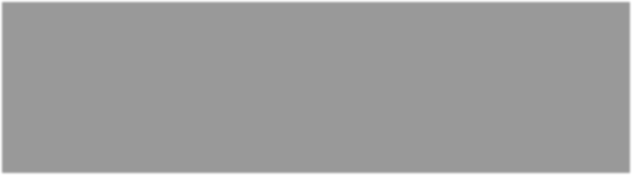

University of Liverpool

website

GP practice

Mumsnet

Netmums

Liverpool Bambis

Liverpool Mums

Facebook

Familiy

member/Friend

Other

*41.*

Where did you hear about this questionnaire?

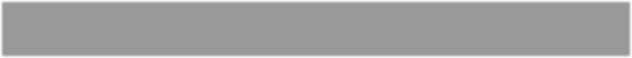

*41*

*.a.*

If you selected Other, please specify:

Page 12

**If you would like to participate further in this study (e.g.	focus groups), please email:**

****

Page 13

***This is the END of the questionnaire.***

**Thank you very much for participating in	this	survey.**

ENTER	THE	PRIZE	DRAW	FOR	A	£100	AMAZON	VOUCHER

**Key	for	selection	options**

**14	- What is your ethnicity?**

White:	British

White:	Irish

White:	Gypsy	or Irish Traveler

White:	Other

Mixed/multiple	ethnic	groups:	White	and Asian

Mixed/multiple	ethnic	groups:	White	and Black African

Mixed/multiple	ethnic	groups:	White	and Black Caribbean

Mixed/multiple	ethnic	groups:	Other

Asian	British/Asian:	Chinese

Asian	British/Asian:	Pakistani

Asian	British/Asian:	Indian

Asian	British/Asian:	Bangladeshi

Asian	British/Asian:	Other

Black	British/Black/African/Caribbean:	African

Black	British/Black/African/Caribbean:	Caribbean

Black	British/Black/African/Caribbean:	Other

Other	ethnic	group:	Arab

Other	ethnic	group:	Other
