## Supplementary file 2. Supplementary images and tables. for "Pregnant women’s attitudes and behaviours towards antenatal vaccination against Influenza and COVID-19 in the Liverpool City Region, United Kingdom: cross-sectional survey"

Image S1. Photo advertisement for questionnaire for distribution through social media outlets.

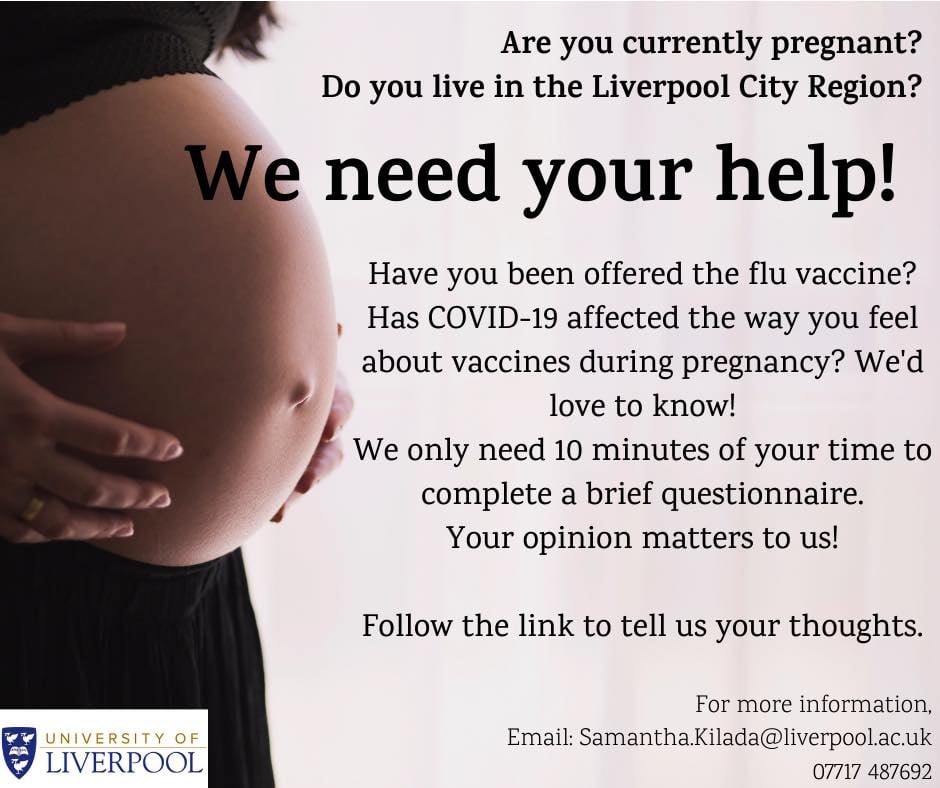

Table S1. Health behaviours for questionnaire respondents in relation to those who were vaccinated/unvaccinated against influenza and those who were accepting, undecided, or against the possibility of the COVID-19 vaccine.

| **Demographic Variables** | **Overall  N=237 (100%)** | **Vaccinated/ intend to against Influenza N=196 (100%)** | **Unvaccinated against Influenza N=39 (100%)** | **Would have COVID-19 Vaccine N=78 (100%)** | **Undecided about having COVID-19 Vaccine N=38 (100%)** | **Would not have COVID-19 Vaccine N=121 (100%)** |
| --- | --- | --- | --- | --- | --- | --- |
| **Pertussis Vaccine** | N=237 | N=196 | N=39 | N=78 | N=38 | N=121 |
| Yes | 177 (74.7%) | 158 (80.6%) | 18.0 (46.2%) | 68.0 (87.2%) | 30.0 (78.9%) | 79.0 (65.3%) |
| No | 60.0 (25.3%) | 38.0 (19.4%) | 21.0 (53.8%) | 10.0 (12.8%) | 8.00 (21.1%) | 42.0 (34.7%) |
| **Antenatal Vitamins** | N=237 | N=196 | N=39 | N=78 | N=38 | N=121 |
| Yes | 206 (86.9%) | 173 (88.3%) | 31.0 (79.5%) | 67.0 (85.9%) | 33.0 (86.8%) | 106 (87.6%) |
| No | 31.0 (13.1%) | 23.0 (11.7%) | 8.00 (20.5%) | 11.0 (14.1%) | 5.00 (13.2%) | 15.0 (12.4%) |
| **Folic Acid** | N=236 | N=195 | N=39 | N=78 | N=38 | N=120 |
| Yes | 192 (81.4%) | 159 (81.5%) | 31.0 (79.5%) | 63.0 (80.8%) | 30.0 (78.9%) | 99.0 (82.5%) |
| No | 44.0 (18.6%) | 36.0 (18.5%) | 8.00 (20.5%) | 15.0 (19.2%) | 8.00 (21.1%) | 21.0 (17.5%) |
| **High Risk** | N=237 | N=196 | N=39 | N=78 | N=38 | N=121 |
| Yes | 35.0 (14.8%) | 32.0 (16.3%) | 3.00 (7.7%) | 12.0 (15.4%) | 5.00 (13.2%) | 18.0 (14.9%) |
| No | 202 (85.2%) | 164 (83.7%) | 36.0 (92.3%) | 66.0 (84.6%) | 33.0 (86.8%) | 103 (85.1%) |
| **Shielded during COVID-19 pandemic** | N=237 | N=196 | N=39 | N=78 | N=38 | N=121 |
| Yes | 45.0 (19.0%) | 38.0 (19.4%) | 7.00 (17.9%) | 15.0 (19.2%) | 8.00 (21.1%) | 22.0 (18.2%) |
| No | 192 (81.0%) | 158 (80.6%) | 32.0 (82.1%) | 63.0 (80.8%) | 30.0 (78.9%) | 99.0 (81.8%) |
| **Days per week exercised before pregnancy** | N=236 | N=195 | N=39 | N=77 | N=38 | N=121 |
| 0 | 17.0 (7.2%) | 14.0 (7.2%) | 3.00 (7.7%) | 4.00 (5.2%) | 0 (0%) | 13.0 (10.7%) |
| 1-2 | 97.0 (41.1%) | 83.0 (42.6%) | 13.0 (33.3%) | 27.0 (35.1%) | 22.0 (57.9%) | 48.0 (39.7%) |
| 3-4 | 77.0 (32.6%) | 58.0 (29.7%) | 18.0 (46.2%) | 28.0 (36.4%) | 9.00 (23.7%) | 40.0 (33.1%) |
| 5-7 | 45.0 (19.1%) | 40.0 (20.5%) | 5.00 (12.8%) | 18.0 (23.4%) | 7.00 (18.4%) | 20.0 (16.5%) |
| **Days per week exercise currently** | N=236 | N=195 | N=39 | N=78 | N=38 | N=120 |
| 0 | 56.0 (23.7%) | 49.0 (25.1%) | 7.00 (17.9%) | 18.0 (23.1%) | 8.00 (21.1%) | 30.0 (25.0%) |
| 1-2 | 106 (44.9%) | 81.0 (41.5%) | 23.0 (59.0%) | 31.0 (39.7%) | 20.0 (52.6%) | 55.0 (45.8%) |
| 3-4 | 49.0 (20.8%) | 43.0 (22.1%) | 6.00 (15.4%) | 19.0 (24.4%) | 7.00 (18.4%) | 23.0 (19.2%) |
| 5-7 | 25.0 (10.6%) | 22.0 (11.3%) | 3.00 (7.7%) | 10.0 (12.8%) | 3.00 (7.9%) | 12.0 (10.0%) |
| **Smoker** | N=234 | N=195 | N=37 | N=77 | N=37 | N=120 |
| Yes | 9.00 (3.8%) | 5.00 (2.6%) | 4.00 (10.8%) | 1.00 (1.3%) | 3.00 (8.1%) | 5.00 (4.2%) |
| No | 225 (96.2%) | 190 (97.4%) | 33.0 (89.2%) | 76.0 (98.7%) | 34.0 (91.9%) | 115 (95.8%) |
| **Discuss vaccination with partner or family member** | N=236 | N=195 | N=39 | N=78 | N=38 | N=120 |
| Yes | 132 (55.9%) | 103 (52.8%) | 28.0 (71.8%) | 41.0 (52.6%) | 19.0 (50.0%) | 72.0 (60.0%) |
| No | 104 (44.0%) | 92.0 (47.2%) | 11.0 (28.2%) | 37.0 (47.4%) | 19.0 (50.0%) | 48.0 (40.0%) |

Table S2. Means by which pregnant women in the Liverpool City Region, UK were offered the influenza vaccine.

| **Questions** | **Overall N=237** |
| --- | --- |
| **Offered flu vaccine** |  |
| Yes | 213 (89.9%) |
| No | 24.0 (10.1%) |
| **Offered flu vaccine by GP** |  |
| Yes | 106 (44.7%) |
| **Offered flu vaccine by Women's Hospital** |  |
| Yes | 15.0 (6.3%) |
| **Offered flu vaccine by community services/midwife** |  |
| Yes | 109 (46.0%) |
| **Offered flu vaccine by health visitor** |  |
| Yes | 7.00 (3.0%) |
| **Offered flu vaccine by pharmacist** |  |
| Yes | 7.00 (3.0%) |
| **Offered flu vaccine by "other"** |  |
| Yes | 39.0 (16.5%) |
| **Offered flu vaccine by "other" specify** |  |
| Employer | 36 (15.2%) |
| Had to request it | 1.00 (0.4%) |
| NHS maternity email updates | 1.00 (0.4%) |
| Whiston Hospital | 1.00 (0.4%) |
| **Offered by letter** |  |
| Yes | 33.0 (13.9%) |
| **Offered by text** |  |
| Yes | 40.0 (16.9%) |
| **Offered face to face** |  |
| Yes | 138 (58.2%) |
| **Offered by other means** |  |
| Yes | 46.0 (19.4%) |
| **Other means specify** |  |
| A leaflet | 1.00 (0.4%) |
| E-mail | 16 (6.8%) |
| Booked/requested themselves | 4 (1.7%) |
| In work on text | 1.00 (0.4%) |
| Intranet from employer | 1.00 (0.4%) |
| Midwife suggestion | 2 (0.8%) |
| Online booking system through trust intranet | 1.00 (0.4%) |
| Telephone | 18 (7.6%) |
| Work | 2.00 (0.8%) |

GP = general practitioner

Table S3. Attitudes and beliefs of pregnant women in the Liverpool City Region, UK towards vaccines in general shown in relation to those who were vaccinated/unvaccinated against influenza and those who were accepting, undecided, or against the possibility of the COVID-19 vaccine.

| **Questions** | **Overall  N=237 (100%)** | **Vaccinated/ intend to against Influenza N=196 (100%)** | **Unvaccinated against Influenza N=39 (100%)** | **Would have COVID-19 Vaccine N=78 (100%)** | **Undecided about having COVID-19 Vaccine N=38 (100%)** | **Would not have COVID-19 Vaccine N=121 (100%)** |
| --- | --- | --- | --- | --- | --- | --- |
| **Vaccines prevent disease.** | N=237 | N=196 | N=39 | N=78 | N=38 | N=121 |
| Disagree | 21.0 (8.9%) | 16.0 (8.2%) | 5.00 (12.8%) | 5.00 (6.4%) | 3.00 (7.9%) | 13.0 (10.7%) |
| Neither Agree or Disagree | 23.0 (9.7%) | 14.0 (7.1%) | 9.00 (23.1%) | 5.00 (6.4%) | 6.00 (15.8%) | 12.0 (9.9%) |
| Agree | 193 (81.4%) | 166 (84.7%) | 25.0 (64.1%) | 68.0 (87.2%) | 29.0 (76.3%) | 96.0 (79.3%) |
| **Vaccines are safe.** | N=237 | N=196 | N=39 | N=78 | N=38 | N=121 |
| Disagree | 3.00 (1.3%) | 1.00 (0.5%) | 2.00 (5.1%) | 0 (0%) | 1.00 (2.6%) | 2.00 (1.7%) |
| Neither Agree or Disagree | 46.0 (19.4%) | 31.0 (15.8%) | 15.0 (38.5%) | 3.00 (3.8%) | 6.00 (15.8%) | 37.0 (30.6%) |
| Agree | 188 (79.3%) | 164 (83.7%) | 22.0 (56.4%) | 75.0 (96.2%) | 31.0 (81.6%) | 82.0 (67.8%) |
| **I intend to vaccinate my child with the flu vaccine when they are old enough.** | N=237 | N=196 | N=39 | N=78 | N=38 | N=121 |
| Disagree | 23.0 (9.7%) | 9.00 (4.6%) | 14.0 (35.9%) | 1.00 (1.3%) | 0 (0%) | 22.0 (18.2%) |
| Neither Agree or Disagree | 26.0 (11.0%) | 14.0 (7.1%) | 12.0 (30.8%) | 3.00 (3.8%) | 5.00 (13.2%) | 18.0 (14.9%) |
| Agree | 188 (79.3%) | 173 (88.3%) | 13.0 (33.3%) | 74.0 (94.9%) | 33.0 (86.8%) | 81.0 (66.9%) |
| **I intend to vaccinate my baby when they are born with all vaccines offered.** | N=237 | N=196 | N=39 | N=78 | N=38 | N=121 |
| Disagree | 3.00 (1.3%) | 0 (0%) | 3.00 (7.7%) | 0 (0%) | 1.00 (2.6%) | 2.00 (1.7%) |
| Neither Agree or Disagree | 7.00 (3.0%) | 2.00 (1.0%) | 5.00 (12.8%) | 1.00 (1.3%) | 0 (0%) | 6.00 (5.0%) |
| Agree | 227 (95.8%) | 194 (99.0%) | 31.0 (79.5%) | 77.0 (98.7%) | 37.0 (97.4%) | 113 (93.4%) |
| **I am more likely to have a vaccine if my family members or friends have had it.** | N=236 | N=196 | N=38 | N=78 | N=37 | N=121 |
| Disagree | 78.0 (33.1%) | 60.0 (30.6%) | 18.0 (47.4%) | 21.0 (26.9%) | 13.0 (35.1%) | 44.0 (36.4%) |
| Neither Agree or Disagree | 77.0 (32.6%) | 66.0 (33.7%) | 10.0 (26.3%) | 33.0 (42.3%) | 8.00 (21.6%) | 36.0 (29.8%) |
| Agree | 81.0 (34.3%) | 70.0 (35.7%) | 10.0 (26.3%) | 24.0 (30.8%) | 16.0 (43.2%) | 41.0 (33.9%) |

Table S4. The likelihood of pregnant women in the Liverpool City Region, UK to accept a vaccine if recommended by different healthcare professionals.

| **I am more likely to have a vaccine if it is recommended by a:** | **Overall N=237 (100%)** | **Vaccinated/ intend to against influenza N=196 (100%)** | **Unvaccinated against influenza N=39 (100%)** |
| --- | --- | --- | --- |
| **Doctor** | N=236 | N=195 | N=39 |
| Disagree | 6.00 (2.5%) | 3.00 (1.5%) | 3.00 (7.7%) |
| Neither Agree or Disagree | 29.0 (12.3%) | 15.0 (7.7%) | 14.0 (35.9%) |
| Agree | 201 (85.2%) | 177 (90.8%) | 22.0 (56.4%) |
| **Pharmacist** | N=231 | N=191 | N=38 |
| Disagree | 21.0 (9.1%) | 15.0 (7.9%) | 6.00 (15.8%) |
| Neither Agree or Disagree | 53.0 (22.9%) | 33.0 (17.3%) | 20.0 (52.6%) |
| Agree | 157 (68.0%) | 143 (74.9%) | 12.0 (31.6%) |
| **Nurse** | N=235 | N=194 | N=39 |
| Disagree | 10.0 (4.3%) | 3.00 (1.5%) | 7.00 (17.9%) |
| Neither Agree or Disagree | 44.0 (18.7%) | 27.0 (13.9%) | 17.0 (43.6%) |
| Agree | 181 (77.0%) | 164 (84.5%) | 15.0 (38.5%) |
| **Midwife** | N=237 | N=196 | N=39 |
| Disagree | 8.00 (3.4%) | 2.00 (1.0%) | 6.00 (15.4%) |
| Neither Agree or Disagree | 29.0 (12.2%) | 15.0 (7.7%) | 14.0 (35.9%) |
| Agree | 200 (84.4%) | 179 (91.3%) | 19.0 (48.7%) |
| **Health visitor** | N=234 | N=193 | N=39 |
| Disagree | 22.0 (9.4%) | 12.0 (6.2%) | 10.0 (25.6%) |
| Neither Agree or Disagree | 48.0 (20.5%) | 29.0 (15.0%) | 19.0 (48.7%) |
| Agree | 164 (70.1%) | 152 (78.8%) | 10.0 (25.6%) |
| **Family member or friend** | N=231 | N=190 | N=39 |
| Disagree | 57.0 (24.7%) | 44.0 (23.2%) | 13.0 (33.3%) |
| Neither Agree or Disagree | 90.0 (39.0%) | 70.0 (36.8%) | 19.0 (48.7%) |
| Agree | 84.0 (36.4%) | 76.0 (40.0%) | 7.00 (17.9%) |

Table S5. The likelihood of pregnant women in the Liverpool City Region, UK to accept the COVID-19 vaccine if recommended by different healthcare professionals.

| **I am more likely to have a COVID-19 vaccine if it is recommended to me by a:** | **Overall N=237 (100%)** | **Would have COVID-19 Vaccine N=78 (100%)** | **Undecided about having COVID-19 Vaccine N=38 (100%)** | **Would not have COVID-19 Vaccine N=121 (100%)** |
| --- | --- | --- | --- | --- |
| **Doctor** | N=236 | N=78 | N=38 | N=120 |
| Disagree | 21.0 (8.9%) | 0 (0%) | 0 (0%) | 21.0 (17.5%) |
| Neither Agree or Disagree | 31.0 (13.1%) | 8.00 (10.3%) | 5.00 (13.2%) | 18.0 (15.0%) |
| Agree | 184 (78.0%) | 70.0 (89.7%) | 33.0 (86.8%) | 81.0 (67.5%) |
| **Pharmacist** | N=228 | N=76 | N=37 | N=115 |
| Disagree | 40.0 (17.5%) | 1.00 (1.3%) | 1.00 (2.7%) | 38.0 (33.0%) |
| Neither Agree or Disagree | 57.0 (25.0%) | 12.0 (15.8%) | 10.0 (27.0%) | 35.0 (30.4%) |
| Agree | 131 (57.5%) | 63.0 (82.9%) | 26.0 (70.3%) | 42.0 (36.5%) |
| **Nurse** | N=230 | N=76 | N=36 | N=110 |
| Disagree | 34.0 (14.3%) | 0 (0%) | 1.00 (2.6%) | 33.0 (27.3%) |
| Neither Agree or Disagree | 43.0 (18.1%) | 10.0 (12.8%) | 9.00 (23.7%) | 24.0 (19.8%) |
| Agree | 153 (64.6%) | 66.0 (84.6%) | 26.0 (68.4%) | 61.0 (50.4%) |
| **Midwife** | N=234 | N=76 | N=38 | N=120 |
| Disagree | 27.0 (11.5%) | 0 (0%) | 0 (0%) | 27.0 (22.5%) |
| Neither Agree or Disagree | 35.0 (15.0%) | 9.00 (11.8%) | 6.00 (15.8%) | 20.0 (16.7%) |
| Agree | 172 (73.5%) | 67.0 (88.1%) | 32.0 (84.2%) | 73.0 (60.8%) |
| **Health visitor** | N=231 | N=74 | N=38 | N=119 |
| Disagree | 42.0 (18.2%) | 2.00 (2.7%) | 4.00 (10.5%) | 36.0 (30.3%) |
| Neither Agree or Disagree | 52.0 (22.5%) | 11.0 (14.9%) | 10.0 (26.3%) | 31.0 (25.8%) |
| Agree | 137 (59.3%) | 61.0 (82.4%) | 24.0 (63.2%) | 52.0 (43.7%) |
| **Family member or friend** | N=227 | N=74 | N=38 | N=115 |
| Disagree | 74.0 (32.6%) | 9.00 (12.2%) | 13.0 (34.2%) | 52.0 (45.2%) |
| Neither Agree or Disagree | 84.0 (37.0%) | 32.0 (43.2%) | 17.0 (44.7%) | 35.0 (30.4%) |
| Agree | 69.0 (30.4%) | 33.0 (44.6%) | 8.00 (21.1%) | 28.0 (24.3%) |
