## Supplementary File 3. R code. for "Pregnant women’s attitudes and behaviours towards antenatal vaccination against Influenza and COVID-19 in the Liverpool City Region, United Kingdom: cross-sectional survey"

Supplementary File 2. R code.

a <- file.choose()

setwd(dirname(a))

require(ggplot2)

require(xlsx)

require(stringr)

require(data.table)

require(ISOweek)

require(ggplot2)

require(epitools)

require(scales)

require(surveillance)

require(Hmisc)

require(dplyr)

require(readxl)

require(stringr)

library(ggpubr)

require(xlsx)

require(finalfit)

require(table1)

require(arsenal)

library(extrafont)

cbPalette <- c("blue","red", "pink", "brown", "black", "green","grey", "orange","yellow","dark red") ##PHE mid color palette

theme_set(theme_bw(base_size = 12, base_family="times"))

coded <- read_excel("questionnaire 1 coded responses.xlsx", sheet = 1, trim_ws = T, col_types = NULL)

coded2<-coded[c("URN","Q1",	"Q2",	"Q3",	"Q4",	"Q5",	"Q6",	"Q6_a",	"Q7",	"Q8",	"Q9",	"Q10",	"Q11_1",	"Q11_2",	"Q11_3",	"Q11_4",	"Q11_a",	"Q12",	"Q13_1",	"Q13_2",	"Q13_3",	"Q13_4",	"Q13_5",	"Q13_6",	"Q14",	"Q15",	"Q16",	"Q17",	"Q18",	"Q19",	"Q22",	"Q22_a_1",	"Q22_a_2",	"Q22_a_3",	"Q22_a_4",	"Q22_a_5",	"Q22_a_6",	"Q22_a_i",	"Q22_b_1",	"Q22_b_2",	"Q22_b_3",	"Q22_b_4",	"Q22_b_i",	"Q23",	"Q23_a",	"Q24",	"Q24_a",	"Q25",	"Q26",	"Q27",	"Q28",	"Q29_1",	"Q29_2",	"Q34",	"Q35",	"Q36",	"Q41_1",	"Q41_2",	"Q41_3",	"Q41_4",	"Q41_5",	"Q41_6",	"Q41_7",	"Q41_8",	"Q41_a",	"CompletionDate")]

noncoded <- read.xlsx("questionnaire 1 non-coded responses.xlsx", sheetName = 1, trim_ws = T)

noncoded2 <- noncoded[c("Unique.Response.Number","X20.1..If.I.get.the.flu..I.will.get.very.ill.",	"X20.2..If.I.get.the.flu..I.will.have.to.stay.home.from.work.school.",	"X20.3..If.I.get.the.flu..my.baby.could.get.ill.",	"X20.4..If.I.get.the.flu..it.could.hurt.my.baby.",	"X20.5..If.I.get.the.flu..my.other.family.members.or.friends.could.get.ill.",	"X20.6..If.I.get.the.flu..my.co.workers.colleagues.could.get.ill.",	"X20.7..If.I.get.the.flu..I.will.die.",	"X21.1..I.feel.knowledgeable.about.the.flu.in.general.",	"X21.2..I.feel.knowledgeable.about.my.risk.of.getting.the.flu.",	"X21.3..I.am.at.risk.of.getting.the.flu.",	"X21.4..My.family.and.friends.are.at.risk.of.getting.the.flu.",	"X30.1..If.I.have.the.flu.vaccine..I.will.have.side.effects.from.it.",	"X30.2..If.I.have.the.flu.vaccine..I.will.get.ill.from.it.",	"X30.3..If.I.have.the.flu.vaccine..it.could.hurt.my.baby.",	"X30.4..If.I.have.the.flu.vaccine..it.will.be.painful.",	"X30.5..If.I.have.the.flu.vaccine..it.will.not.protect.me.from.getting.the.flu.",	"X30.6..If.I.have.the.flu.vaccine..it.will.not.protect.my.baby.",	"X30.a.1..It.is.inconvenient.for.me.to.get.the.flu.vaccine.",	"X30.a.2..There.is.a.shortage.of.the.flu.vaccine.",	"X30.a.3..The.flu.vaccine.was.recommended.to.me.by.my.healthcare.provider..e.g..doctor..nurse..midwife..",	"X31.1..If.I.have.the.flu.vaccine..I.will.not.get.ill.with.the.flu.",	"X31.2..If.I.have.the.flu.vaccine..I.will.help.prevent.my.baby.from.getting.the.flu.",	"X31.3..If.I.have.the.flu.vaccine..I.will.help.prevent.my.family.friends.from.getting.ill.with.the.flu.",	"X32.1..Vaccines.prevent.disease.",	"X32.2..Vaccines.are.safe.",	"X32.3..I.intend.to.vaccinate.my.child.with.the.flu.vaccine.when.they.are.old.enough.",	"X32.4..I.intend.to.vaccinate.my.baby.when.they.are.born.with.all.vaccines.offered.",	"X32.5..I.am.more.likely.to.have.a.vaccine.if.my.family.members.or.friends.have.had.it.",	"X33.1....doctor",	"X33.2....pharmacist",	"X33.3....nurse",	"X33.4....midwife",	"X33.5....health.visitor",	"X33.6....family.member.or.friend",	"X37.1..If.I.get.COVID.19..I.will.get.very.ill.",	"X37.2..If.I.get.COVID.19..I.will.have.to.isolate.myself.",	"X37.3..If.I.get.COVID.19..my.family.members.and.friends.who.came.in.contact.with.me.will.have.to.quarantine.themselves.",	"X37.4..If.I.get.COVID.19..my.baby.could.get.ill.",	"X37.5..If.I.get.COVID.19..my.other.family.members.or.friends.could.get.ill.",	"X38.1..I.feel.knowledgeable.about.COVID.19.in.general.",	"X38.2..I.feel.knowledgeable.about.my.risk.of.getting.COVID.19.",	"X38.3..I.am.at.risk.of.getting.COVID.19.",	"X38.4..My.family.and.friends.are.at.risk.of.getting.COVID.19",	"X39.1..If.a.COVID.19.vaccine.was.available.to.me.now..I.would.get.it.",	"X39.2..A.COVID.19.vaccine.would.protect.me.",	"X39.3..A.COVID.19.vaccine.would.protect.my.baby..other.family.members..or.friends.from.getting.ill.with.COVID.19.",	"X39.4..I.would.vaccinate.my.baby.against.COVID.19.as.soon.as.possible.after.they.are.born.",	"X39.5..If.a.COVID.19.vaccine.was.seasonal..I.would.get.it.every.year.",	"X39.6..I.would.get.a.COVID.19.vaccine.if.I.wasn.t.pregnant.",	"X40.1....doctor.",	"X40.2....pharmacist.",	"X40.3....nurse.",	"X40.4....midwife.",	"X40.5....health.visitor.",	"X40.6....family.member.or.friend")]

noncoded[noncoded==""]<-NA

full<-cbind(coded2,noncoded2)

#rename columns

colnames(full)

names <- c("URN",	"Consent",	"Age",	"Weeks.pregnant",	"No.of.children",	"High.risk",	"Education.Lvl",	"Education.other",	"Occupation",	"Postcode",	"Income",	"Smartphone",	"Access.GP.online.appts",	"Access.GP.online.prescrp",	"Access.GP.online.records",	"Access.GP.online.other",	"Access.other.specify",	"No.in.household",	"Live.with.alone",	"Live.with.partner",	"Live.with.child.ren",	"Live.with.parent.s",	"Live.with.other.family",	"Live.with.roommate.s",	"Ethnicity",	"Smoker",	"Exercise.before",	"Exercise.currently",	"Antenatal.vitamins",	"Folic.acid",	"Offered.flu.vacc",	"If.yes.offered.GP",	"If.yes.offered.wo.hosp",	"If.yes.offered.comm.ser.midwife",	"If.yes.offered.health.vis",	"If.yes.offered.pharm",	"If.yes.offered.other",	"Who.other.offered.specify",	"How.offered.letter",	"How.offered.text",	"How.offered.face.to.face",	"How.offered.other",	"How.other.offered.specify",	"Flu.vacc.this.preg",	"If.no.intend",	"Flu.vacc.prev.preg",	"If.yes.side.effects",	"Discuss.vacc.with.partner.fam",	"You.or.close.friend.fam.flu",	"OTC.meds.flu",	"Alt.med.flu",	"Pertuss.vacc.yes",	"Pertuss.vacc.no",	"You.or.close.friend.fam.pos.COVID",	"You.or.close.friend.fam.hosp.COVID",	"Shield",	"Heard.Uni.site",	"Heard.GP.prac",	"Heard.Mumsnet",	"Heard.Netmums",	"Heard.Bambis",	"Heard.Liv.Mums",	"Heard.fam.friend",	"Heard.other",	"Heard.ques.Other.specify",	"Completion.Date","Unique.Response.Number",	"Flu.ill",	"Flu.stay.home.work.school",	"Flu.baby.ill",	"Flu.hurt.baby",	"Flu.family.friends.ill",	"Flu.coworkers.colleages.ill",	"Flu.die",	"Knowledgeable.flu.general",	"Knowledgeable.flu.risk",	"I.am.risk.getting.flu",	"Family.friends.risk.getting.flu",	"Flu.vacc.side.effects",	"Flu.vacc.ill",	"Flu.vacc.hurt.baby",	"Flu.vacc.painful",	"Flu.vacc.not.protect.me",	"Flu.vacc.not.protect.baby",	"Inconvenient.flu.vacc",	"Shortage.flu.vacc",	"Flu.vacc.recommended",	"Flu.vacc.not.get.ill",	"Flu.vacc.prevent.baby.ill",	"Flu.vacc.prevent.family.friends.ill",	"Vacc.prevent.disease",	"Vacc.safe",	"Intend.vacc.child.flu",	"Intend.vacc.baby",	"More.likely.vacc.if.family.friends.have",	"Doc.flu",	"Pharm.flu",	"Nurse.flu",	"Midwife.flu",	"Health.visitor.flu",	"Fam.friend.flu",	"COVID.ill",	"COVID.isolate",	"COVID.friends",	"COVID.could.contact.quarantine",	"COVID.family.friends.contact.quarantine",	"Knowledge.COVID.general",	"Knowledge.COVID.risk",	"I.am.risk.getting.COVID",	"Family.friends.risk.getting.COVID",	"If.COVID.vacc.available.would.have",	"COVID.vacc.protect.me",	"COVID.vacc.protect.baby.fam.friends",	"Would.vacc.baby.COVID",	"If.COVID.vacc.seasonal.would.have",	"Would.have.COVID.vacc.if.not.preg",	"Doc.COVID",	"Pharm.COVID",	"Nurse.COVID",	"Midwife.COVID",	"Health.visitor.COVID",	"Fam.friend.COVID")

colnames(full) <- names

#set date fields

full$Completion.Date <- as.Date(full$Completion.Date, origin = "1899-12-30")

full$yearmonth<-format(full$Completion.Date,"%Y%m")

#Pre post April 16th

full$PostApril16<-0

full$PostApril16[full$Completion.Date>="2021-04-16"]<-1

#completion date

full$monthresponse<-format(full$Completion.Date, '%B')

full$monthresponse<-as.factor(full$monthresponse)

#check number of rows

nrow(full)

#remove outliers for age

full$Age <- as.numeric(full$Age)

full1 <- subset(full, Age<70&!is.na(Age))

#postcodes

full1$Postcode<-substr(full1$Postcode, 1, 4)

full1$Postcode<-toupper(full1$Postcode)

full1 <- subset(full1, subset=(Postcode!="CH7" & Postcode!="WN6" & Postcode!="CH66" & Postcode!="WA1" & Postcode!="CW7" & Postcode!="CW1" & Postcode!="PR5" & Postcode!="GU34" & Postcode!="HU6"))

#relabel income

full1$Income<-as.factor(full1$Income)

levels(full1$Income)<-c("<£10,000", "£10,001-20,000", "£20,001-30,000","£30,001-45,000","£45,001-60,000",">£60,000")

##create a new field for income and recode

full1$Income2<-as.character(full1$Income)

full1$Income2[full1$Income2 =="<£10,000" | full1$Income2 =="£10,001-20,000" | full1$Income2 =="£20,001-30,000"]<-"<=£30,000"

full1$Income2[full1$Income =="£30,001-45,000" |full1$Income =="£45,001-60,000"]<-"£30,001-60,000"

full1$Income2<-as.factor(full1$Income2)

levels(full1$Income2)

full1$Income2<-factor(full1$Income2,levels(full1$Income2)[c(2,1,3)])

levels(full1$Income2)

#recode occupation

full1$Occupation2<-NA

full1$Occupation2[full1$Occupation=="Property manager"| full1$Occupation==	"Research manager"| full1$Occupation==	"Dentist"| full1$Occupation==	"veterinary surgeon"| full1$Occupation==	"Self employed"| full1$Occupation==	"GP trainee"| full1$Occupation==	"Project Manager"| full1$Occupation==	"Quality Systems Analyst"| full1$Occupation==	"Head of Year (pastoral role in secondary education)"| full1$Occupation==	"Marketing Manager"| full1$Occupation==	"Data analyst"| full1$Occupation==	"Civil Engineer"| full1$Occupation==	"Policy Manager for NHS England"| full1$Occupation==	"Critical Care Outreach Practitioner"| full1$Occupation==	"Town planner"| full1$Occupation==	"Doctor"| full1$Occupation==	"Property manager"| full1$Occupation==	"Research and Finance"| full1$Occupation==	"Scientist"| full1$Occupation==	"NHS Senior Improvement Advisor"| full1$Occupation==	"Clinical psychologist"| full1$Occupation==	"Recruitment consultant"| full1$Occupation==	"Fe lecturer"| full1$Occupation==	"Lecturer & researcher"| full1$Occupation==	"Portfolio Research Facilitator"| full1$Occupation==	"Solicitor"| full1$Occupation==	"Hospital dentist"| full1$Occupation==	"Self-employed"| full1$Occupation==	"Project coordinator"| full1$Occupation==	"Quality Assessor"| full1$Occupation== "Business analyst"| full1$Occupation==	"Manager at Manufacturing Facility"| full1$Occupation==	"Chartered accountant"| full1$Occupation==	"Recruitment Manager"| full1$Occupation==	"Quality administration"| full1$Occupation==	"Business Analyst"| full1$Occupation==	"Head teacher"| full1$Occupation==	"Change analyst"| full1$Occupation==	"Sales Co-ordinator"| full1$Occupation==	"Project Manager"| full1$Occupation==	"Quality specialist"| full1$Occupation==	"Business Analyst"| full1$Occupation==	"Pharmacist "]<- "More advantaged groups NS-SEC groups 1-4"

full1$Occupation2[full1$Occupation==	"Clinical Psychologist"| full1$Occupation==	"Senior Researcher (Charity sector)"| full1$Occupation==	"Anaesthetist"| full1$Occupation==	"Pharmacist"| full1$Occupation==	"Vet and research scientist"| full1$Occupation==	"Lecturer"| full1$Occupation==	"Marketing Manager"| full1$Occupation==	"Solicitor"| full1$Occupation==	"Director tax and accounting"| full1$Occupation==	"Self employed"| full1$Occupation==	"Lead HR Business Partner"| full1$Occupation==	"Pharmacist "| full1$Occupation==	"Veterinary Surgeon"| full1$Occupation=="Project manager"| full1$Occupation==	"Urban Design Officer"| full1$Occupation== "Local Government"| full1$Occupation==	"Lecturer university"| full1$Occupation==	"Doctor (GP)"| full1$Occupation==	"Compaby Director"| full1$Occupation==	"Project Manager"| full1$Occupation==	"Speech and Language Therapist"| full1$Occupation==	"Management Consultant"| full1$Occupation==	"Hospital Doctor"| full1$Occupation==	"Nurse"| full1$Occupation==	"Events manager"| full1$Occupation==	"Physiotherapist"| full1$Occupation==	"Investment Banker"| full1$Occupation==	"Teacher"| full1$Occupation==	"Teacher"| full1$Occupation==	"Lab technician"| full1$Occupation==	"Neonatal ANP"| full1$Occupation==	"Nhs Health Care Assistant"| full1$Occupation==	"Teacher"| full1$Occupation==	"Occupational therapy"| full1$Occupation==	"Teacher"| full1$Occupation==	"Business Development Coordinator for Charitable Orgs "| full1$Occupation==	"Community Heart Failure Nurse"| full1$Occupation==	"Nurse"| full1$Occupation==	"Librarian"| full1$Occupation=="Laboratory tech"| full1$Occupation==	"Veterinary Surgeon"| full1$Occupation==	"Social worker "| full1$Occupation==	"Behaviour Analyst"| full1$Occupation==	"ABA therapist"| full1$Occupation==	"Educator"| full1$Occupation==	"Supply teacher"| full1$Occupation==	"Community Engagement Officer"| full1$Occupation==	"Physiotherapist"| full1$Occupation==	"Marketing "| full1$Occupation==	"Staff Trainer"| full1$Occupation==	"Account Director"| full1$Occupation==	"mental health nurse"| full1$Occupation==	"Communications Officer"| full1$Occupation==	"Teacher"| full1$Occupation==	"Nurse"| full1$Occupation==	"Special needs teacher"| full1$Occupation==	"Physiotherapist"| full1$Occupation==	"NHS Personal Assistant"]<- "More advantaged groups NS-SEC groups 1-4"

full1$Occupation2[full1$Occupation==	"Registered nurse"| full1$Occupation==	"NHS"| full1$Occupation==	"Communications Officer "| full1$Occupation==	"District Nurse"| full1$Occupation==	"Account Manager"| full1$Occupation==	"Staff Nurse"| full1$Occupation==	"Radiotherapist"| full1$Occupation=="Account Manager"| full1$Occupation==	"Education "| full1$Occupation==	"Illustrator"| full1$Occupation==	"student nurse"| full1$Occupation==	"Mental health nurse"| full1$Occupation==	"Letting agent"| full1$Occupation==	"Nurse"| full1$Occupation==	"Finance assistant"| full1$Occupation==	"Marketer"| full1$Occupation==	"Healthcare assistant"| full1$Occupation==	"Care assistant"| full1$Occupation==	"Teacher"| full1$Occupation==	"Therapy Radiographer"| full1$Occupation==	"Manager/social worker"| full1$Occupation==	"Marketing officer"| full1$Occupation==	"Registered Nurse"| full1$Occupation==	"Technician"| full1$Occupation==	"Teacher"| full1$Occupation==	"Mental health nurse"| full1$Occupation==	"Teacher"| full1$Occupation==	"Copywriter"| full1$Occupation==	"Manager"| full1$Occupation==	"Nurse"| full1$Occupation==	"Supervisor retail"| full1$Occupation==	"Student nurse "| full1$Occupation==	"Teacher"| full1$Occupation==	"MIS Officer"| full1$Occupation==	"Specialist Nurse"| full1$Occupation==	"Care assistant"| full1$Occupation==	"Customer services manager"| full1$Occupation==	"Financial Advisor "| full1$Occupation==	"Nurse"| full1$Occupation==	"Therapy assistant"| full1$Occupation=="Nursery deputy manager"| full1$Occupation==	"Nurse"| full1$Occupation==	"Learning disability nurse"| full1$Occupation==	"Video Producer"| full1$Occupation==	"Healthcare assistant"| full1$Occupation==	"Process Technologist (Food Manufacturing)"| full1$Occupation==	"Nurse"| full1$Occupation==	"Teacher"| full1$Occupation==	"Business development manager "| full1$Occupation==	"Manager"| full1$Occupation==	"Training Officer"| full1$Occupation==	"Teacher"| full1$Occupation==	"Nurse"| full1$Occupation==	"Nursery worker"| full1$Occupation==	"Student Midwife"| full1$Occupation==	"Housing officer"| full1$Occupation==	"Nurse"| full1$Occupation==	"Registered nurse"| full1$Occupation==	"Business Development Coordinator"| full1$Occupation==	"Student Money Adviser"| full1$Occupation==	"Senior Healthcare Assistant"| full1$Occupation==	"Teacher"| full1$Occupation==	"Care coordinator NHS"| full1$Occupation==	"Research assistant"| full1$Occupation==	"Bank manager"] <- "More advantaged groups NS-SEC groups 1-4"

full1$Occupation2[full1$Occupation==	"Supply teacher"| full1$Occupation==	"Physiotherapist"| full1$Occupation==	"SEN teacher "| full1$Occupation=="Healthcare assistant"| full1$Occupation==	"Supervisor at an Investment Bank"| full1$Occupation==	"Looked After Children`s Nurse"| full1$Occupation==	"Police officer"| full1$Occupation==	"Agency"| full1$Occupation==	"Administration"| full1$Occupation==	"Local goverment officer"| full1$Occupation==	"Finance Administrator"| full1$Occupation==	"Client account administration"| full1$Occupation==	"Operations Coordinator"| full1$Occupation==	"Civil servant"| full1$Occupation==	"Executive Assistant"| full1$Occupation==	"Bank clerk"| full1$Occupation==	"Administrator"| full1$Occupation==	"Administrator"| full1$Occupation==	"Police Officer"| full1$Occupation==	"Care assistant"| full1$Occupation==	"HR admin"| full1$Occupation==	"Teaching assistant "| full1$Occupation==	"Early years practitioner"| full1$Occupation==	"Admin role"| full1$Occupation==	"Customer servic advisor"| full1$Occupation==	"Training Administrator"| full1$Occupation==	"Office admin"| full1$Occupation==	"Nursery nurse"| full1$Occupation==	"Childcare practitioner"| full1$Occupation=="Call center operator"| full1$Occupation==	"Teaching Assistant"| full1$Occupation==	"Civil servant"| full1$Occupation==	"Office staff"| full1$Occupation==	"Administrator in the NHS"| full1$Occupation==	"Customer care"| full1$Occupation==	"Nursery nurse"| full1$Occupation==	"Call centre"| full1$Occupation==	"Teaching assistant"| full1$Occupation==	"Claims Handler"| full1$Occupation==	"Police Staff"| full1$Occupation==	"Marine administrator"| full1$Occupation==	"business banker"| full1$Occupation==	"Special Needs Teaching Assistant"| full1$Occupation==	"Designer"| full1$Occupation==	"Licensee"] <- "More advantaged groups NS-SEC groups 1-4"

full1$Occupation2[is.na(full1$Occupation2)] <- "Less advantaged groups NS-SEC groups 5-8"

full1$Occupation2[full1$Occupation=="Student" |full1$Occupation==	"Dog Walker and student" |is.na(full1$Occupation) |full1$Occupation=="Postgraduate student"]<-"Exceptions"

#reorder SEC

full1$Occupation2<-as.factor(full1$Occupation2)

levels(full1$Occupation2)

full1$Occupation2<-factor(full1$Occupation2,levels(full1$Occupation2)[c(3,2,1)])

levels(full1$Occupation2)

#Create a new field for ethnicity

full1$Ethnicity2<-"BAME"

full1$Ethnicity2[full1$Ethnicity=="1"]<-"White British"

full1$Ethnicity2[full1$Ethnicity=="2" | full1$Ethnicity=="3" | full1$Ethnicity=="4"]<-"White Other"

#reorder ethnicity

full1$Ethnicity2<-as.factor(full1$Ethnicity2)

levels(full1$Ethnicity2)

full1$Ethnicity2<-factor(full1$Ethnicity2,levels(full1$Ethnicity2)[c(2,3,1)])

levels(full1$Ethnicity2)

#create new field for number of children

full1$No.of.children2<-NA

full1$No.of.children2[full1$No.of.children=="1"]<-"This is my first"

full1$No.of.children2[full1$No.of.children=="2"]<-"1"

full1$No.of.children2[full1$No.of.children=="3"]<-"2"

full1$No.of.children2[full1$No.of.children=="4" | full1$No.of.children=="5"]<-"3+"

#reorder number of children

full1$No.of.children2<-as.factor(full1$No.of.children2)

levels(full1$No.of.children2)

full1$No.of.children2<-factor(full1$No.of.children2,levels(full1$No.of.children2)[c(4,1,2,3)])

levels(full1$No.of.children2)

#create new field for number in household

full1$No.in.household2<-NA

full1$No.in.household2[full1$No.in.household=="1"]<-"1"

full1$No.in.household2[full1$No.in.household=="2"]<-"2"

full1$No.in.household2[full1$No.in.household=="3"]<-"3"

full1$No.in.household2[full1$No.in.household=="4" | full1$No.in.household=="5" | full1$No.in.household=="8"]<-"4+"

#put weeks pregnant in numeric

full1$Weeks.pregnant<-as.numeric(full1$Weeks.pregnant)

#pertussis

full1$Pertussis<-NA

full1$Pertussis[full1$Pertuss.vacc.yes=="1"]<-"Yes"

full1$Pertussis[full1$Pertuss.vacc.no=="1"]<-"No"

#reorder pertussis

full1$Pertussis<-as.factor(full1$Pertussis)

levels(full1$Pertussis)

full1$Pertussis<-factor(full1$Pertussis,levels(full1$Pertussis)[c(2,1)])

levels(full1$Pertussis)

#recode education

full1$Education2<-NA

full1$Education2[full1$Education.Lvl=="1"]<-"Below GCSE"

full1$Education2[full1$Education.Lvl=="2"]<-"GCSE or similar"

full1$Education2[full1$Education.Lvl=="3"]<-"NVQ or similar"

full1$Education2[full1$Education.Lvl=="4"]<-"A-level or similar"

full1$Education2[full1$Education.Lvl=="5"]<-"Undergraduate"

full1$Education2[full1$Education.Lvl=="6"]<-"Postgraduate"

full1$Education2[full1$Education.Lvl=="7"]<-"Other"

#reorder education

full1$Education2<-as.factor(full1$Education2)

levels(full1$Education2)

full1$Education2<-factor(full1$Education2,levels(full1$Education2)[c(2,3,1,6,5,4)])

levels(full1$Education2)

#put high risk in numeric

##full1$High.risk<-as.numeric(full1$High.risk)

#new flu vaccine field

full1$final.flu.vacc.this.Preg <- full1$Flu.vacc.this.preg

full1$final.flu.vacc.this.Preg[full1$Flu.vacc.this.preg=="2" & full1$If.no.intend=="1"] <- "1"

table(full1$flu.vacc.this.preg,full1$If.no.intend)

#recode OTC medicine

full1$OTC.meds.flu2<-NA

full1$OTC.meds.flu2[full1$OTC.meds.flu=="1"]<-"Yes"

full1$OTC.meds.flu2[full1$OTC.meds.flu=="2"]<-"No"

#reorder OTC medicine

full1$OTC.meds.flu2<-as.factor(full1$OTC.meds.flu2)

levels(full1$OTC.meds.flu2)

full1$OTC.meds.flu2<-factor(full1$OTC.meds.flu2,levels(full1$OTC.meds.flu2)[c(2,1)])

levels(full1$OTC.meds.flu2)

#recode alternative flu medicine

full1$Alt.med.flu2<-NA

full1$Alt.med.flu2[full1$Alt.med.flu=="1"]<-"Yes"

full1$Alt.med.flu2[full1$Alt.med.flu=="2"]<-"No"

#reorder alternative flu medicine

full1$Alt.med.flu2<-as.factor(full1$Alt.med.flu2)

levels(full1$Alt.med.flu2)

full1$Alt.med.flu2<-factor(full1$Alt.med.flu2,levels(full1$Alt.med.flu2)[c(2,1)])

levels(full1$Alt.med.flu2)

#recode antenatal vitamins

full1$Antenatal.vitamins2<-NA

full1$Antenatal.vitamins2[full1$Antenatal.vitamins=="1"]<-"Yes"

full1$Antenatal.vitamins2[full1$Antenatal.vitamins=="2"]<-"No"

#reorder antenatal vitamins

full1$Antenatal.vitamins2<-as.factor(full1$Antenatal.vitamins2)

levels(full1$Antenatal.vitamins2)

full1$Antenatal.vitamins2<-factor(full1$Antenatal.vitamins2,levels(full1$Antenatal.vitamins2)[c(2,1)])

levels(full1$Antenatal.vitamins2)

#recode folic acid

full1$Folic.acid2<-NA

full1$Folic.acid2[full1$Folic.acid=="1"]<-"Yes"

full1$Folic.acid2[full1$Folic.acid=="2"]<-"No"

#reorder folic acid

full1$Folic.acid2<-as.factor(full1$Folic.acid2)

levels(full1$Folic.acid2)

full1$Folic.acid2<-factor(full1$Folic.acid2,levels(full1$Folic.acid2)[c(2,1)])

levels(full1$Folic.acid2)

#reorder if COVID vaccine available would you have it

full1$If.COVID.vacc.available.would.have<-as.factor(full1$If.COVID.vacc.available.would.have)

levels(full1$If.COVID.vacc.available.would.have)

full1$If.COVID.vacc.available.would.have<-factor(full1$If.COVID.vacc.available.would.have,levels(full1$If.COVID.vacc.available.would.have)[c(5,2,3,1,4)])

levels(full1$If.COVID.vacc.available.would.have)

#recode likert scales

full1<-full1 %>%

mutate_at(c(68:122), funs(recode(., `Strongly Disagree`="Disagree", `Disagree`="Disagree", `Neither Agree or Disagree`="Neither Agree or Disagree", `Agree`="Agree", `Strongly Agree`="Agree", .default = as.character(NA))))

#recode high risk

full1$High.risk2<-NA

full1$High.risk2[full1$High.risk=="1"]<-"Yes"

full1$High.risk2[full1$High.risk=="2"]<-"No"

#reorder high risk

full1$High.risk2<-as.factor(full1$High.risk2)

levels(full1$High.risk2)

full1$High.risk2<-factor(full1$High.risk2,levels(full1$High.risk2)[c(2,1)])

levels(full1$High.risk2)

###reorder likert questions###

full1$Flu.ill<-as.factor(full1$Flu.ill)

levels(full1$Flu.ill)

full1$Flu.ill<-factor(full1$Flu.ill,levels(full1$Flu.ill)[c(2,3,1)])

levels(full1$Flu.ill)

#_______________________________________________________________________________________________________

full1$Flu.stay.home.work.school<-as.factor(full1$Flu.stay)

levels(full1$Flu.stay.home.work.school)

full1$Flu.stay.home.work.school<-factor(full1$Flu.stay.home.work.school,levels(full1$Flu.stay.home.work.school)[c(2,3,1)])

levels(full1$Flu.stay.home.work.school)

#_______________________________________________________________________________________________________

full1$Flu.baby.ill<-as.factor(full1$Flu.baby.ill)

levels(full1$Flu.baby.ill)

full1$Flu.baby.ill<-factor(full1$Flu.baby.ill,levels(full1$Flu.baby.ill)[c(2,3,1)])

levels(full1$Flu.baby.ill)

#_______________________________________________________________________________________________________

full1$Flu.hurt.baby<-as.factor(full1$Flu.hurt.baby)

levels(full1$Flu.hurt.baby)

full1$Flu.hurt.baby<-factor(full1$Flu.hurt.baby,levels(full1$Flu.hurt.baby)[c(2,3,1)])

levels(full1$Flu.hurt.baby)

#_______________________________________________________________________________________________________

full1$Flu.family.friends.ill<-as.factor(full1$Flu.family.friends.ill)

levels(full1$Flu.family.friends.ill)

full1$Flu.family.friends.ill<-factor(full1$Flu.family.friends.ill,levels(full1$Flu.family.friends.ill)[c(2,3,1)])

levels(full1$Flu.family.friends.ill)

#_______________________________________________________________________________________________________

full1$Flu.coworkers.colleages.ill<-as.factor(full1$Flu.coworkers.colleages.ill)

levels(full1$Flu.coworkers.colleages.ill)

full1$Flu.coworkers.colleages.ill<-factor(full1$Flu.coworkers.colleages.ill,levels(full1$Flu.coworkers.colleages.ill)[c(2,3,1)])

levels(full1$Flu.coworkers.colleages.ill)

#_______________________________________________________________________________________________________

full1$Flu.die<-as.factor(full1$Flu.die)

levels(full1$Flu.die)

full1$Flu.die<-factor(full1$Flu.die,levels(full1$Flu.die)[c(2,3,1)])

levels(full1$Flu.die)

#_______________________________________________________________________________________________________

full1$Knowledgeable.flu.general<-as.factor(full1$Knowledgeable.flu.general)

levels(full1$Knowledgeable.flu.general)

full1$Knowledgeable.flu.general<-factor(full1$Knowledgeable.flu.general,levels(full1$Knowledgeable.flu.general)[c(2,3,1)])

levels(full1$Knowledgeable.flu.general)

#_______________________________________________________________________________________________________

full1$Knowledgeable.flu.risk<-as.factor(full1$Knowledgeable.flu.risk)

levels(full1$Knowledgeable.flu.risk)

full1$Knowledgeable.flu.risk<-factor(full1$Knowledgeable.flu.risk,levels(full1$Knowledgeable.flu.risk)[c(2,3,1)])

levels(full1$Knowledgeable.flu.risk)

#_______________________________________________________________________________________________________

full1$I.am.risk.getting.flu<-as.factor(full1$I.am.risk.getting.flu)

levels(full1$I.am.risk.getting.flu)

full1$I.am.risk.getting.flu<-factor(full1$I.am.risk.getting.flu,levels(full1$I.am.risk.getting.flu)[c(2,3,1)])

levels(full1$I.am.risk.getting.flu)

#_______________________________________________________________________________________________________

full1$Family.friends.risk.getting.flu<-as.factor(full1$Family.friends.risk.getting.flu)

levels(full1$Family.friends.risk.getting.flu)

full1$Family.friends.risk.getting.flu<-factor(full1$Family.friends.risk.getting.flu,levels(full1$Family.friends.risk.getting.flu)[c(2,3,1)])

levels(full1$Family.friends.risk.getting.flu)

#_______________________________________________________________________________________________________

full1$Flu.vacc.side.effects<-as.factor(full1$Flu.vacc.side.effects)

levels(full1$Flu.vacc.side.effects)

full1$Flu.vacc.side.effects<-factor(full1$Flu.vacc.side.effects,levels(full1$Flu.vacc.side.effects)[c(2,3,1)])

levels(full1$Flu.vacc.side.effects)

#_______________________________________________________________________________________________________

full1$Flu.vacc.ill<-as.factor(full1$Flu.vacc.ill)

levels(full1$Flu.vacc.ill)

full1$Flu.vacc.ill<-factor(full1$Flu.vacc.ill,levels(full1$Flu.vacc.ill)[c(2,3,1)])

levels(full1$Flu.vacc.ill)

#_______________________________________________________________________________________________________

full1$Flu.vacc.hurt.baby<-as.factor(full1$Flu.vacc.hurt.baby)

levels(full1$Flu.vacc.hurt.baby)

full1$Flu.vacc.hurt.baby<-factor(full1$Flu.vacc.hurt.baby,levels(full1$Flu.vacc.hurt.baby)[c(2,3,1)])

levels(full1$Flu.vacc.hurt.baby)

#_______________________________________________________________________________________________________

full1$Flu.vacc.painful<-as.factor(full1$Flu.vacc.painful)

levels(full1$Flu.vacc.painful)

full1$Flu.vacc.painful<-factor(full1$Flu.vacc.painful,levels(full1$Flu.vacc.painful)[c(2,3,1)])

levels(full1$Flu.vacc.painful)

#_______________________________________________________________________________________________________

full1$Flu.vacc.not.protect.me<-as.factor(full1$Flu.vacc.not.protect.me)

levels(full1$Flu.vacc.not.protect.me)

full1$Flu.vacc.not.protect.me<-factor(full1$Flu.vacc.not.protect.me,levels(full1$Flu.vacc.not.protect.me)[c(2,3,1)])

levels(full1$Flu.vacc.not.protect.me)

#_______________________________________________________________________________________________________

full1$Flu.vacc.not.protect.baby<-as.factor(full1$Flu.vacc.not.protect.baby)

levels(full1$Flu.vacc.not.protect.baby)

full1$Flu.vacc.not.protect.baby<-factor(full1$Flu.vacc.not.protect.baby,levels(full1$Flu.vacc.not.protect.baby)[c(2,3,1)])

levels(full1$Flu.vacc.not.protect.baby)

#_______________________________________________________________________________________________________

full1$Inconvenient.flu.vacc<-as.factor(full1$Inconvenient.flu.vacc)

levels(full1$Inconvenient.flu.vacc)

full1$Inconvenient.flu.vacc<-factor(full1$Inconvenient.flu.vacc,levels(full1$Inconvenient.flu.vacc)[c(2,3,1)])

levels(full1$Inconvenient.flu.vacc)

#_______________________________________________________________________________________________________

full1$Shortage.flu.vacc<-as.factor(full1$Shortage.flu.vacc)

levels(full1$Shortage.flu.vacc)

full1$Shortage.flu.vacc<-factor(full1$Shortage.flu.vacc,levels(full1$Shortage.flu.vacc)[c(2,3,1)])

levels(full1$Shortage.flu.vacc)

#_______________________________________________________________________________________________________

full1$Flu.vacc.recommended<-as.factor(full1$Flu.vacc.recommended)

levels(full1$Flu.vacc.recommended)

full1$Flu.vacc.recommended<-factor(full1$Flu.vacc.recommended,levels(full1$Flu.vacc.recommended)[c(2,3,1)])

levels(full1$Flu.vacc.recommended)

#_______________________________________________________________________________________________________

full1$Flu.vacc.not.get.ill<-as.factor(full1$Flu.vacc.not.get.ill)

levels(full1$Flu.vacc.not.get.ill)

full1$Flu.vacc.not.get.ill<-factor(full1$Flu.vacc.not.get.ill,levels(full1$Flu.vacc.not.get.ill)[c(2,3,1)])

levels(full1$Flu.vacc.not.get.ill)

#_______________________________________________________________________________________________________

full1$Flu.vacc.prevent.baby.ill<-as.factor(full1$Flu.vacc.prevent.baby.ill)

levels(full1$Flu.vacc.prevent.baby.ill)

full1$Flu.vacc.prevent.baby.ill<-factor(full1$Flu.vacc.prevent.baby.ill,levels(full1$Flu.vacc.prevent.baby.ill)[c(2,3,1)])

levels(full1$Flu.vacc.prevent.baby.ill)

#_______________________________________________________________________________________________________

full1$Flu.vacc.prevent.family.friends.ill<-as.factor(full1$Flu.vacc.prevent.family.friends.ill)

levels(full1$Flu.vacc.prevent.family.friends.ill)

full1$Flu.vacc.prevent.family.friends.ill<-factor(full1$Flu.vacc.prevent.family.friends.ill,levels(full1$Flu.vacc.prevent.family.friends.ill)[c(2,3,1)])

levels(full1$Flu.vacc.prevent.family.friends.ill)

#_______________________________________________________________________________________________________

full1$Vacc.prevent.disease<-as.factor(full1$Vacc.prevent.disease)

levels(full1$Vacc.prevent.disease)

full1$Vacc.prevent.disease<-factor(full1$Vacc.prevent.disease,levels(full1$Vacc.prevent.disease)[c(2,3,1)])

levels(full1$Vacc.prevent.disease)

#_______________________________________________________________________________________________________

full1$Vacc.safe<-as.factor(full1$Vacc.safe)

levels(full1$Vacc.safe)

full1$Vacc.safe<-factor(full1$Vacc.safe,levels(full1$Vacc.safe)[c(2,3,1)])

levels(full1$Vacc.safe)

#_______________________________________________________________________________________________________

full1$Intend.vacc.child.flu<-as.factor(full1$Intend.vacc.child.flu)

levels(full1$Intend.vacc.child.flu)

full1$Intend.vacc.child.flu<-factor(full1$Intend.vacc.child.flu,levels(full1$Intend.vacc.child.flu)[c(2,3,1)])

levels(full1$Intend.vacc.child.flu)

#_______________________________________________________________________________________________________

full1$Intend.vacc.baby<-as.factor(full1$Intend.vacc.baby)

levels(full1$Intend.vacc.baby)

full1$Intend.vacc.baby<-factor(full1$Intend.vacc.baby,levels(full1$Intend.vacc.baby)[c(2,3,1)])

levels(full1$Intend.vacc.baby)

#_______________________________________________________________________________________________________

full1$More.likely.vacc.if.family.friends.have<-as.factor(full1$More.likely.vacc.if.family.friends.have)

levels(full1$More.likely.vacc.if.family.friends.have)

full1$More.likely.vacc.if.family.friends.have<-factor(full1$More.likely.vacc.if.family.friends.have,levels(full1$More.likely.vacc.if.family.friends.have)[c(2,3,1)])

levels(full1$More.likely.vacc.if.family.friends.have)

#_______________________________________________________________________________________________________

full1$Doc.flu<-as.factor(full1$Doc.flu)

levels(full1$Doc.flu)

full1$Doc.flu<-factor(full1$Doc.flu,levels(full1$Doc.flu)[c(2,3,1)])

levels(full1$Doc.flu)

#_______________________________________________________________________________________________________

full1$Pharm.flu<-as.factor(full1$Pharm.flu)

levels(full1$Pharm.flu)

full1$Pharm.flu<-factor(full1$Pharm.flu,levels(full1$Pharm.flu)[c(2,3,1)])

levels(full1$Pharm.flu)

#_______________________________________________________________________________________________________

full1$Nurse.flu<-as.factor(full1$Nurse.flu)

levels(full1$Nurse.flu)

full1$Nurse.flu<-factor(full1$Nurse.flu,levels(full1$Nurse.flu)[c(2,3,1)])

levels(full1$Nurse.flu)

#_______________________________________________________________________________________________________

full1$Midwife.flu<-as.factor(full1$Midwife.flu)

levels(full1$Midwife.flu)

full1$Midwife.flu<-factor(full1$Midwife.flu,levels(full1$Midwife.flu)[c(2,3,1)])

levels(full1$Midwife.flu)

#_______________________________________________________________________________________________________

full1$Health.visitor.flu<-as.factor(full1$Health.visitor.flu)

levels(full1$Health.visitor.flu)

full1$Health.visitor.flu<-factor(full1$Health.visitor.flu,levels(full1$Health.visitor.flu)[c(2,3,1)])

levels(full1$Health.visitor.flu)

#_______________________________________________________________________________________________________

full1$Fam.friend.flu<-as.factor(full1$Fam.friend.flu)

levels(full1$Fam.friend.flu)

full1$Fam.friend.flu<-factor(full1$Fam.friend.flu,levels(full1$Fam.friend.flu)[c(2,3,1)])

levels(full1$Fam.friend.flu)

#_______________________________________________________________________________________________________

full1$COVID.ill<-as.factor(full1$COVID.ill)

levels(full1$COVID.ill)

full1$COVID.ill<-factor(full1$COVID.ill,levels(full1$COVID.ill)[c(2,3,1)])

levels(full1$COVID.ill)

#_______________________________________________________________________________________________________

full1$COVID.isolate<-as.factor(full1$COVID.isolate)

levels(full1$COVID.isolate)

full1$COVID.isolate<-factor(full1$COVID.isolate,levels(full1$COVID.isolate)[c(2,3,1)])

levels(full1$COVID.isolate)

#_______________________________________________________________________________________________________

full1$COVID.friends<-as.factor(full1$COVID.friends)

levels(full1$COVID.friends)

full1$COVID.friends<-factor(full1$COVID.friends,levels(full1$COVID.friends)[c(2,3,1)])

levels(full1$COVID.friends)

#_______________________________________________________________________________________________________

full1$COVID.could.contact.quarantine<-as.factor(full1$COVID.could.contact.quarantine)

levels(full1$COVID.could.contact.quarantine)

full1$COVID.could.contact.quarantine<-factor(full1$COVID.could.contact.quarantine,levels(full1$COVID.could.contact.quarantine)[c(2,3,1)])

levels(full1$COVID.could.contact.quarantine)

#_______________________________________________________________________________________________________

full1$COVID.family.friends.contact.quarantine<-as.factor(full1$COVID.family.friends.contact.quarantine)

levels(full1$COVID.family.friends.contact.quarantine)

full1$COVID.family.friends.contact.quarantine<-factor(full1$COVID.family.friends.contact.quarantine,levels(full1$COVID.family.friends.contact.quarantine)[c(2,3,1)])

levels(full1$COVID.family.friends.contact.quarantine)

#_______________________________________________________________________________________________________

full1$Knowledge.COVID.general<-as.factor(full1$Knowledge.COVID.general)

levels(full1$Knowledge.COVID.general)

full1$Knowledge.COVID.general<-factor(full1$Knowledge.COVID.general,levels(full1$Knowledge.COVID.general)[c(2,3,1)])

levels(full1$Knowledge.COVID.general)

#_______________________________________________________________________________________________________

full1$Knowledge.COVID.risk<-as.factor(full1$Knowledge.COVID.risk)

levels(full1$Knowledge.COVID.risk)

full1$Knowledge.COVID.risk<-factor(full1$Knowledge.COVID.risk,levels(full1$Knowledge.COVID.risk)[c(2,3,1)])

levels(full1$Knowledge.COVID.risk)

#_______________________________________________________________________________________________________

full1$I.am.risk.getting.COVID<-as.factor(full1$I.am.risk.getting.COVID)

levels(full1$I.am.risk.getting.COVID)

full1$I.am.risk.getting.COVID<-factor(full1$I.am.risk.getting.COVID,levels(full1$I.am.risk.getting.COVID)[c(2,3,1)])

levels(full1$I.am.risk.getting.COVID)

#_______________________________________________________________________________________________________

full1$Family.friends.risk.getting.COVID<-as.factor(full1$Family.friends.risk.getting.COVID)

levels(full1$Family.friends.risk.getting.COVID)

full1$Family.friends.risk.getting.COVID<-factor(full1$Family.friends.risk.getting.COVID,levels(full1$Family.friends.risk.getting.COVID)[c(2,3,1)])

levels(full1$Family.friends.risk.getting.COVID)

#_______________________________________________________________________________________________________

full1$COVID.vacc.protect.me<-as.factor(full1$COVID.vacc.protect.me)

levels(full1$COVID.vacc.protect.me)

full1$COVID.vacc.protect.me<-factor(full1$COVID.vacc.protect.me,levels(full1$COVID.vacc.protect.me)[c(2,3,1)])

levels(full1$COVID.vacc.protect.me)

#_______________________________________________________________________________________________________

full1$COVID.vacc.protect.baby.fam.friends<-as.factor(full1$COVID.vacc.protect.baby.fam.friends)

levels(full1$COVID.vacc.protect.baby.fam.friends)

full1$COVID.vacc.protect.baby.fam.friends<-factor(full1$COVID.vacc.protect.baby.fam.friends,levels(full1$COVID.vacc.protect.baby.fam.friends)[c(2,3,1)])

levels(full1$COVID.vacc.protect.baby.fam.friends)

#_______________________________________________________________________________________________________

full1$Would.vacc.baby.COVID<-as.factor(full1$Would.vacc.baby.COVID)

levels(full1$Would.vacc.baby.COVID)

full1$Would.vacc.baby.COVID<-factor(full1$Would.vacc.baby.COVID,levels(full1$Would.vacc.baby.COVID)[c(2,3,1)])

levels(full1$Would.vacc.baby.COVID)

#_______________________________________________________________________________________________________

full1$If.COVID.vacc.seasonal.would.have<-as.factor(full1$If.COVID.vacc.seasonal.would.have)

levels(full1$If.COVID.vacc.seasonal.would.have)

full1$If.COVID.vacc.seasonal.would.have<-factor(full1$If.COVID.vacc.seasonal.would.have,levels(full1$If.COVID.vacc.seasonal.would.have)[c(2,3,1)])

levels(full1$If.COVID.vacc.seasonal.would.have)

#_______________________________________________________________________________________________________

full1$Would.have.COVID.vacc.if.not.preg<-as.factor(full1$Would.have.COVID.vacc.if.not.preg)

levels(full1$Would.have.COVID.vacc.if.not.preg)

full1$Would.have.COVID.vacc.if.not.preg<-factor(full1$Would.have.COVID.vacc.if.not.preg,levels(full1$Would.have.COVID.vacc.if.not.preg)[c(2,3,1)])

levels(full1$Would.have.COVID.vacc.if.not.preg)

#_______________________________________________________________________________________________________

full1$Doc.COVID<-as.factor(full1$Doc.COVID)

levels(full1$Doc.COVID)

full1$Doc.COVID<-factor(full1$Doc.COVID,levels(full1$Doc.COVID)[c(2,3,1)])

levels(full1$Doc.COVID)

#_______________________________________________________________________________________________________

full1$Pharm.COVID<-as.factor(full1$Pharm.COVID)

levels(full1$Pharm.COVID)

full1$Pharm.COVID<-factor(full1$Pharm.COVID,levels(full1$Pharm.COVID)[c(2,3,1)])

levels(full1$Pharm.COVID)

#_______________________________________________________________________________________________________

full1$Nurse.COVID<-as.factor(full1$Nurse.COVID)

levels(full1$Nurse.COVID)

full1$Nurse.COVID<-factor(full1$Nurse.COVID,levels(full1$Nurse.COVID)[c(2,3,1)])

levels(full1$Nurse.COVID)

#_______________________________________________________________________________________________________

full1$Midwife.COVID<-as.factor(full1$Midwife.COVID)

levels(full1$Midwife.COVID)

full1$Midwife.COVID<-factor(full1$Midwife.COVID,levels(full1$Midwife.COVID)[c(2,3,1)])

levels(full1$Midwife.COVID)

#_______________________________________________________________________________________________________

full1$Health.visitor.COVID<-as.factor(full1$Health.visitor.COVID)

levels(full1$Health.visitor.COVID)

full1$Health.visitor.COVID<-factor(full1$Health.visitor.COVID,levels(full1$Health.visitor.COVID)[c(2,3,1)])

levels(full1$Health.visitor.COVID)

#_______________________________________________________________________________________________________

full1$Fam.friend.COVID<-as.factor(full1$Fam.friend.COVID)

levels(full1$Fam.friend.COVID)

full1$Fam.friend.COVID<-factor(full1$Fam.friend.COVID,levels(full1$Fam.friend.COVID)[c(2,3,1)])

levels(full1$Fam.friend.COVID)

#recode flu vacc to include those who intend

full1$fluyes<-NA

full1$fluyes[full1$Flu.vacc.this.preg==1]<-"1"

full1$fluyes[full1$Flu.vacc.this.preg==2]<-"2"

full1$fluyes[full1$If.no.intend==1]<-"1"

full1$fluyes[full1$If.no.intend==2]<-"2"

full1$fluyes[full1$Flu.vacc.this.preg==2 & is.na(full1$If.no.intend)]<-NA

#reorder month respons

levels(full1$monthresponse)

full1$monthresponse<-factor(full1$monthresponse,levels(full1$monthresponse)[c(6,2,4,3,5,1)])

levels(full1$monthresponse)

#######################################descriptive tables##############################################

table1(~ Age + Occupation2 +Income+ Income2+Ethnicity+ Ethnicity2+No.of.children2+No.of.children+No.in.household+No.in.household2+OTC.meds.flu2+Alt.med.flu2+Antenatal.vitamins2+Folic.acid2+Pertussis+Education.Lvl+Education2 +If.COVID.vacc.available.would.have, data=full1)

table1(~ Age + Occupation2 +Income+ Income2+Ethnicity+ Ethnicity2+No.of.children2+No.of.children+No.in.household+No.in.household2+OTC.meds.flu2+Alt.med.flu2+Antenatal.vitamins2+Folic.acid2+Pertussis+Education.Lvl+Education2+If.no.intend| Flu.vacc.this.preg, data=full1)

table1(~ Age + Occupation2 +Income+ Income2+ Ethnicity2+No.of.children2+No.in.household2+OTC.meds.flu2+Alt.med.flu2+Antenatal.vitamins2+Folic.acid2+Pertussis+Education2+If.no.intend+Flu.vacc.this.preg| If.COVID.vacc.available.would.have, data=full1)

summary(Flu.vacc.this.preg ~ Age + Occupation2 + Income2 + Education2 + Ethnicity2 + No.of.children2 + No.in.household2 + Pertussis + Antenatal.vitamins2 + High.risk2 + OTC.meds.flu2 + Alt.med.flu2, data=full1, method="reverse", test=T)

summary( If.COVID.vacc.available.would.have ~ Age + Occupation2 +Income+ Income2+ Ethnicity2+No.of.children2+No.in.household2+OTC.meds.flu+Alt.med.flu+Antenatal.vitamins+Folic.acid+Pertussis+Education2+If.no.intend+Flu.vacc.this.preg, data=full1, method="reverse", test=T)

summary(If.COVID.vacc.available.would.have ~ Age + Occupation2 + Income2+ Ethnicity2+No.of.children2+No.in.household2+OTC.meds.flu2+Alt.med.flu2+Antenatal.vitamins+Pertussis+Education2+If.no.intend+Flu.vacc.this.preg + High.risk2, data=full1, method="reverse", test=T)

table1(~ Age + Occupation2 + Income2 + Ethnicity2 + No.of.children2 + No.in.household2 | Flu.vacc.prev.preg, data=full1)

#_______

table1(~ Age + Weeks.pregnant + Occupation2 + Income2 + Education2 + Ethnicity2 + No.of.children2 + No.in.household2 + Pertussis + Antenatal.vitamins2 + Folic.acid2 + High.risk2 + Shield + Smartphone + Exercise.before + Exercise.currently + Smoker + Discuss.vacc.with.partner.fam + monthresponse + as.factor(PostApril16)| fluyes, data=full1)

table1(~ Age + Weeks.pregnant + Occupation2 + Income2 + Education2 + Ethnicity2 + No.of.children2 + No.in.household2 + Pertussis + Antenatal.vitamins2 + Folic.acid2 + High.risk2 + Shield + Smartphone + Exercise.before + Exercise.currently + Smoker + Discuss.vacc.with.partner.fam + monthresponse + as.factor(PostApril16)| If.COVID.vacc.available.would.have, data=full1)

table1(~ Flu.ill + Flu.stay.home.work.school + Flu.baby.ill + Flu.hurt.baby+ Flu.family.friends.ill + Flu.coworkers.colleages.ill + Flu.die + Knowledgeable.flu.general + Knowledgeable.flu.risk + I.am.risk.getting.flu + Family.friends.risk.getting.flu + You.or.close.friend.fam.flu + OTC.meds.flu2 + Alt.med.flu2| fluyes, data=full1)

table1(~ Flu.vacc.side.effects + Flu.vacc.ill + Flu.vacc.hurt.baby + Flu.vacc.painful + Flu.vacc.not.protect.me + Flu.vacc.not.protect.baby + Inconvenient.flu.vacc + Shortage.flu.vacc + Flu.vacc.recommended + Flu.vacc.not.get.ill + Flu.vacc.prevent.baby.ill + Flu.vacc.prevent.family.friends.ill +Flu.vacc.prev.preg + If.yes.side.effects | fluyes, data=full1)

table1(~ Vacc.prevent.disease + Vacc.safe + Intend.vacc.child.flu + Intend.vacc.baby + More.likely.vacc.if.family.friends.have | fluyes, data=full1)

table1(~ Vacc.prevent.disease + Vacc.safe + Intend.vacc.child.flu + Intend.vacc.baby + More.likely.vacc.if.family.friends.have | If.COVID.vacc.available.would.have, data=full1)

table1(~ Doc.flu + Pharm.flu + Nurse.flu + Midwife.flu + Health.visitor.flu + Fam.friend.flu | fluyes, data=full1)

table1(~ COVID.ill + COVID.isolate + COVID.friends + COVID.could.contact.quarantine + COVID.family.friends.contact.quarantine + Knowledge.COVID.general + Knowledge.COVID.general + Knowledge.COVID.risk + I.am.risk.getting.COVID + Family.friends.risk.getting.COVID + You.or.close.friend.fam.pos.COVID + You.or.close.friend.fam.hosp.COVID | If.COVID.vacc.available.would.have, data=full1)

table1(~ If.COVID.vacc.available.would.have + COVID.vacc.protect.me + COVID.vacc.protect.baby.fam.friends + Would.vacc.baby.COVID + If.COVID.vacc.seasonal.would.have + Would.have.COVID.vacc.if.not.preg, data=full1)

table1(~ Doc.COVID + Pharm.COVID + Nurse.COVID + Midwife.COVID + Health.visitor.COVID + Fam.friend.COVID | If.COVID.vacc.available.would.have, data=full1)

table1(~ Offered.flu.vacc + If.yes.offered.GP + If.yes.offered.wo.hosp + If.yes.offered.comm.ser.midwife + If.yes.offered.health.vis + If.yes.offered.pharm + If.yes.offered.other + Who.other.offered.specify + How.offered.letter + How.offered.text + How.offered.face.to.face + How.offered.other + How.other.offered.specify, data=full1)

#table1(~ Access.GP.online.appts + Access.GP.online.prescrp + Access.GP.online.records + Access.GP.online.other + Live.with.alone + Live.with.partner + Live.with.child.ren + Live.with.parent.s + Live.with.other.family + Live.with.roommate.s, data=full1)

#table1(~ Heard.Uni.site + Heard.GP.prac + Heard.Mumsnet + Heard.Netmums + Heard.Bambis + Heard.Liv.Mums + Heard.fam.friend + Heard.other + Heard.ques.Other.specify, data=full1)

table1(~ Flu.vacc.this.preg + If.no.intend + Flu.vacc.prev.preg + If.yes.side.effects, data=full1)

#Don`t use this

#require(rmarkdown)

#table_one <- tableby(formula = as.factor(Flu.vacc.this.preg) ~ Age+Income2+fe(Ethnicity2), data = full1,control = tableby.control(numeric.stats="medianq1q3"))

#summary(table_one)

#write2html(table_one, "~/trash.html")

#write2word(table_one, "~/trash.doc", title="My table in Word")

table1(~ If.COVID.vacc.available.would.have | You.or.close.friend.fam.pos.COVID, data=full1)

table1(~ If.COVID.vacc.available.would.have | You.or.close.friend.fam.hosp.COVID, data=full1)

table1(~ If.COVID.vacc.available.would.have | fluyes, data=full1)

table1(~ fluyes | You.or.close.friend.fam.flu, data=full1)

tab1<-tableby(fluyes ~ Age + Weeks.pregnant, data=full1, control=tableby.control( numeric.stats=c("N", "median", "q1q3")))

tab2<-tableby(If.COVID.vacc.available.would.have ~ Age + Weeks.pregnant, data=full1, control=tableby.control( numeric.stats=c("N", "median", "q1q3")))

###################################### descriptive plots ##############################################

hist(full1$Age, main = "", xlab = "Age (weeks)")

mean(full1$Age)

min(full1$Age)

max(full1$Age)

hist(full1$Weeks.pregnant, main = "", xlab = "Weeks Pregnant")

hist(full1$High.risk)

summary(full1$Age)

IQR(full1$Age)

summary(full1$Weeks.pregnant)

IQR(full1$Weeks.pregnant)
